# The adverse impact of COVID-19 pandemic on cardiovascular disease prevention and management in England, Scotland and Wales: A population-scale analysis of trends in medication data

**DOI:** 10.1101/2021.12.31.21268587

**Authors:** Caroline E Dale, Rohan Takhar, Ray Carragher, Fatemeh Torabi, Michalis Katsoulis, Stephen Duffield, Seamus Kent, Tanja Mueller, Amanj Kurdi, Stuart McTaggart, Hoda Abbasizanjani, Sam Hollings, Andrew Scourfield, Ronan Lyons, Rowena Griffiths, Jane Lyons, Gareth Davies, Dan Harris, Alex Handy, Mehrdad Alizadeh Mizani, Chris Tomlinson, Mark Ashworth, Spiros Denaxas, Amitava Banerjee, Jonathan Sterne, Kate Lovibond, Paul Brown, Ian Bullard, Rouven Priedon, Mamas A Mamas, Ann Slee, Paula Lorgelly, Munir Pirmohamed, Kamlesh Khunti, Naveed Sattar, Andrew Morris, Cathie Sudlow, Ashley Akbari, Marion Bennie, Reecha Sofat, on behalf of the CVD-COVID-UK Consortium

**Author notes:** Joint first authors (Caroline E Dale, Rohan Takhar).

## Abstract

**Objectives:** To estimate the impact of the COVID-19 pandemic on cardiovascular disease (CVD) and CVD management using routinely collected medication data as a proxy.

**Design:** Descriptive and interrupted time series analysis using anonymised individual-level population-scale data for 1.32 billion records of dispensed CVD medications across 15.8 million individuals in England, Scotland and Wales.

**Setting:** Community dispensed CVD medications with 100% coverage from England, Scotland and Wales, plus primary care prescribed CVD medications from England (including 98% English general practices).

**Participants:** 15.8 million individuals aged 18+ years alive on 1^st^ April 2018 dispensed at least one CVD medicine in a year from England, Scotland and Wales.

**Main outcome measures:** Monthly counts, percent annual change (1^st^ April 2018 to 31^st^ July 2021) and annual rates (1^st^ March 2018 to 28^th^ February 2021) of medicines dispensed by CVD/ CVD risk factor; prevalent and incident use.

**Results:** Year-on-year change in dispensed CVD medicines by month were observed, with notable uplifts ahead of the first (11.8% higher in March 2020) but not subsequent national lockdowns. Using hypertension as one example of the indirect impact of the pandemic, we observed 491,203 fewer individuals initiated antihypertensive treatment across England, Scotland and Wales during the period March 2020 to end May 2021 than would have been expected compared to 2019. We estimated that this missed antihypertension treatment could result in 13,659 additional CVD events should individuals remain untreated, including 2,281 additional myocardial infarctions (MIs) and 3,474 additional strokes. Incident use of lipid-lowering medicines decreased by an average 14,793 per month in early 2021 compared with the equivalent months prior to the pandemic in 2019. In contrast, the use of incident medicines to treat type-2 diabetes (T2DM) increased by approximately 1,642 patients per month.

**Conclusions:** Management of key CVD risk factors as proxied by incident use of CVD medicines has not returned to pre-pandemic levels in the UK. Novel methods to identify and treat individuals who have missed treatment are urgently required to avoid large numbers of additional future CVD events, further adding indirect cost of the COVID-19 pandemic.

## Introduction

Cardiovascular disease (CVD) remains the most common cause of mortality and morbidity worldwide; it is therefore vital to understand the impact of the COVID-19 pandemic on CVD and its risk factors. In the UK, strategies for CVD prevention include screening for health conditions and risk factors that can be modified through medication including Type-2 diabetes (T2DM), hypertension, hypercholesterolaemia and atrial fibrillation (AF). When adequately controlled, such measures reduce the level of CVD in the population.

The COVID-19 pandemic has disrupted health care in multiple ways, putting additional pressure on both primary and secondary care services^1-3^. How these have impacted on screening and treatment of common risk factors, including CVD risk factors, and the downstream impact of missed detection of incident disease remains under studied at a national level^4^.

Medicines are a key public health intervention, particularly in preventing and controlling long-term health conditions. Examining the change in the prescribed and dispensed medicines used to treat CVD risk factors over the course of the COVID-19 pandemic can be used to model and assess the likely impact of not treating these risk factors on future CVD events This is a complementary strategy to studying reduction in the level of disease diagnoses and risk factor control; and maybe a closer representation of the real-world control of CVD risk factors within the population, following the logical patient pathway from diagnosis, through prescription, to dispensing of medication to the treatment of the condition. In this study, we investigate the impact of COVID-19 on non-COVID harm, specifically the management of CVD in eleven sub-populations defined by medicines. We report, for the first time across >60M people in England, Scotland and Wales how a medicines-based treatment approach can provide precise and comprehensive quantification of the reduction in the control of CVD risk factors due to the pandemic.

## Methods

### Data

We studied anonymised individual-level population-scale data from England, Scotland and Wales accessed through the respective national Trusted Research Environments (TREs), i.e. NHS Digital’s TRE for England (referred to throughout as ‘the English TRE’), the Scottish National Safe Haven and the SAIL Databank. Motivated by the public health importance of understanding the relationship between COVID-19 and CVD, the Health Data Research UK (HDR UK) British Heart Foundation (BHF) Data Science Centre (DSC) established the CVD-COVID-UK consortium and related research programme^5,6^. Through this partnership, linked, nationally-collated electronic health record (EHR) data for the population of England, Scotland and Wales have been made available to support research into the impacts of CVD on COVID-19 and vice versa. Details of the collaboration and the data included within each of the national TREs are described in full elsewhere. Data processing for this work are available under an open source license at https://github.com/BHFDSC/CCU014_01.

#### England

Medication data are available from a number of sources within the English TRE. First, prescribed data are available within the General Practice Extraction Service (GPES) extract Data for Pandemic Planning and Research (GDPPR), including data from 98% of all English general practices. These medicines were *a priori* selected by the CVD-COVID-UK programme predominently for their relevance to CVD and its risk factors (e.g. cholesterol lowering, antiplatelet/ anticoagulant, antihypertensive, diabetes)^7^. These prescribed data include the exact date of prescription for each drug item. Second, the NHS Business Service Authority (NHSBSA) dispensing data are updated on a monthly basis and include prescriptions for all medicines dispensesd in the community in England^8^. The English TRE also provides data for some secondary care Electronic Prescribing and Medicines Administration (EPMA), but these are not included in the current analyses as most medicines to prevent and treat CVD are mainly accounted for by primary care prescribing.

#### Scotland

The medicines data available within the Scottish National Safe Haven^9,10^ come from the Prescribing Information System (PIS), which captures all prescriptions dispensed in the community in Scotland^11^. In the main, these prescriptions originate from general practitioners, although other health care professionals (e.g. dentists, pharmacists) may also issue prescriptions. PIS uses a drug categorisation system based on the British National Formulary (BNF) with the majority of the data coming through community pharmacies via the Data Capture Validation Pricing (DCVP) system. Both paper and electronic prescriptions are provided as part of Scotland’s eHealth strategy. The data is updated monthly on the Safe Haven. The exact date of prescribing is available in PIS; but since PIS only captures prescriptions that have been dispensed the focus here is on Scottish dispensing data.

#### Wales

Primary care prescribing and dispensing data for the population of Wales are available from two main data sources within the Secure Anonymised Information Linkage (SAIL) Databank^12,13^. Firstly, prescribing data from approximately 80% of all Wales general practices are available within the Welsh Longitudinal General Practice (WLGP) data, which is updated on a monthly basis^14^. These data include the exact date of prescription for each drug item and are coded using Read codes. Secondly, dispensing data from all community pharmacies in Wales is available within the Welsh Dispensing Data Set (WDDS)^15^, which is updated on a monthly basis. Within SAIL upon each monthly release of WDDS, a research ready data asset (RRDA) is created and maintained^16^ based on COVID-19 population e-cohort RRDA^12^, which enhances the dispensing data for research purposes with mapping to additional coding classifications and meta-data. Although primary care prescribing data is available for the population of Wales, a comprehensive mapping between Read and BNF codes is currenty not available. Therefore, in these analyses we have focused only on Welsh dispensing data.

### Categorisation of CVD risk factor medications

Medicines were selected from BNF Chapters 2 (Cardiovascular System) and 6 (Endocrine System)^17^. These were manually curated (initially by RS; reviewed by AS) to choose therapies used to treat and/ or licenced to treat the major CVD risk factors: blood pressure, AF, LDL-cholesterol and diabetes. Anticoagulants were categorised on their own and by class: vitamin K antagonists (VKA), direct oral anticoagulants (DOAC) and heparins. This allowed analysis of behaviours within the anticoagulant category; for example, differential use of VKAs and DOACs during the pandemic. Some medicines are used in more than one category; for example, anticoagulants may be used to treat AF (most commonly VKAs and DOACs) but can also be used to treat deep vein thrombosis and pulmonary embolism. Medicines used to treat hypertension and heart failure are also often overlapping. Here medicines were categorised and analysed in only one group to prevent double counting. Hence most blood pressure lowering agents were classified as antihypertensives apart from some classes of beta blockers, loop diuretics (and some thiazides e.g. metolazone), and sacubitril/valsartan which are used specifically for heart failure. This may result in undercounting for medicines used in heart failure in these analyses and consequently heart failure as a condition may be under-represented. Antiplatelets were classified on their own as they can be used for primary and secondary prevention for MI, stroke and peripheral vascular disease (PVD). An additional and separate category of medicines that are mainly used as anti-anginals was created. Insulins and other glucose lowering therapies for T2DM were categorised separately, although it is well known that a proportion of individuals with T2DM will also be on insulins. However, understanding the trends over time in the separate categories may be important specifically in tracking incident T2DM. Like antihypertensives certain medicines may have more than one indication, e.g. SGLT-2 inhbitors are now licenced for heart failure. Additional analyses could be carried out linking to disease code although this was out of scope for the analyses presesented here. Excluded medicines were all intravenous preparations, those used to treat pulmonary hypertension, anti-arrhythmics where the indication is unlikely to be AF, sclerosants and medicines with very low prescription rates. Medicines included in each of the 11 CVD sub-groups: Antihypertensives, Antiplatelets secondary prevention (primary for DM), DOAC, Warfarin, Heparins, Lipid lowering, T2DM, Insulin, Heart failure, AF, Angina are shown in **Appendix Table 1**.

### Medication Data Processing

A detailed description of the medications data processing undertaken in each national TRE is given in the **Supplementary Methods** and at https://github.com/BHFDSC/CCU014_01. For all analyses (with the exception of the interrupted time series analysis), dispensing data were used as these are more likely to be indicative of individuals taking medicines and were available in all three nations. Within the English TRE, NHSBSA was screened to identify all possible dispensed medicines. Both dispensed and prescribed data were mapped to the British National Formulary (BNF)^17^ (via DM+D or SNOMED concepts), and the medication substance identified using the 8^th^ BNF character to facilitate categorisation according to CVD sub-group.

Analyses were mirrored in the Scottish National Safe Haven & SAIL Databank using the same inclusion criteria, code lists and categorisation for CVD medicines using BNF codes selected and extracted from the English TRE, with some adjustments required due to specifics of the datasets in each (see **Supplementary Methods** for description in full). Summary output files from each nation were extracted and combined with results from other nations.

In this study we focused on the major modifiable cardiovascular risk factors with antihypertensives, lipid-lowering medications, T2DM and insulin medications. We presented results for dispensed medications for these four CVD sub-groups combined and by each individual sub-group. Results on medications for the remaining seven CVD sub-groups are presented in the **Supplementary Material**. Within each of the CVD sub-groups, we identified the first time a medication was dispensed to an individual during the study period (**Figure 1**). By highlighting monthly trends in first (incident) medication use, we can understand changes in the rate of identification of CVD risk factors and unmet control within individuals due to the pandemic.

**FIGURE 1:**
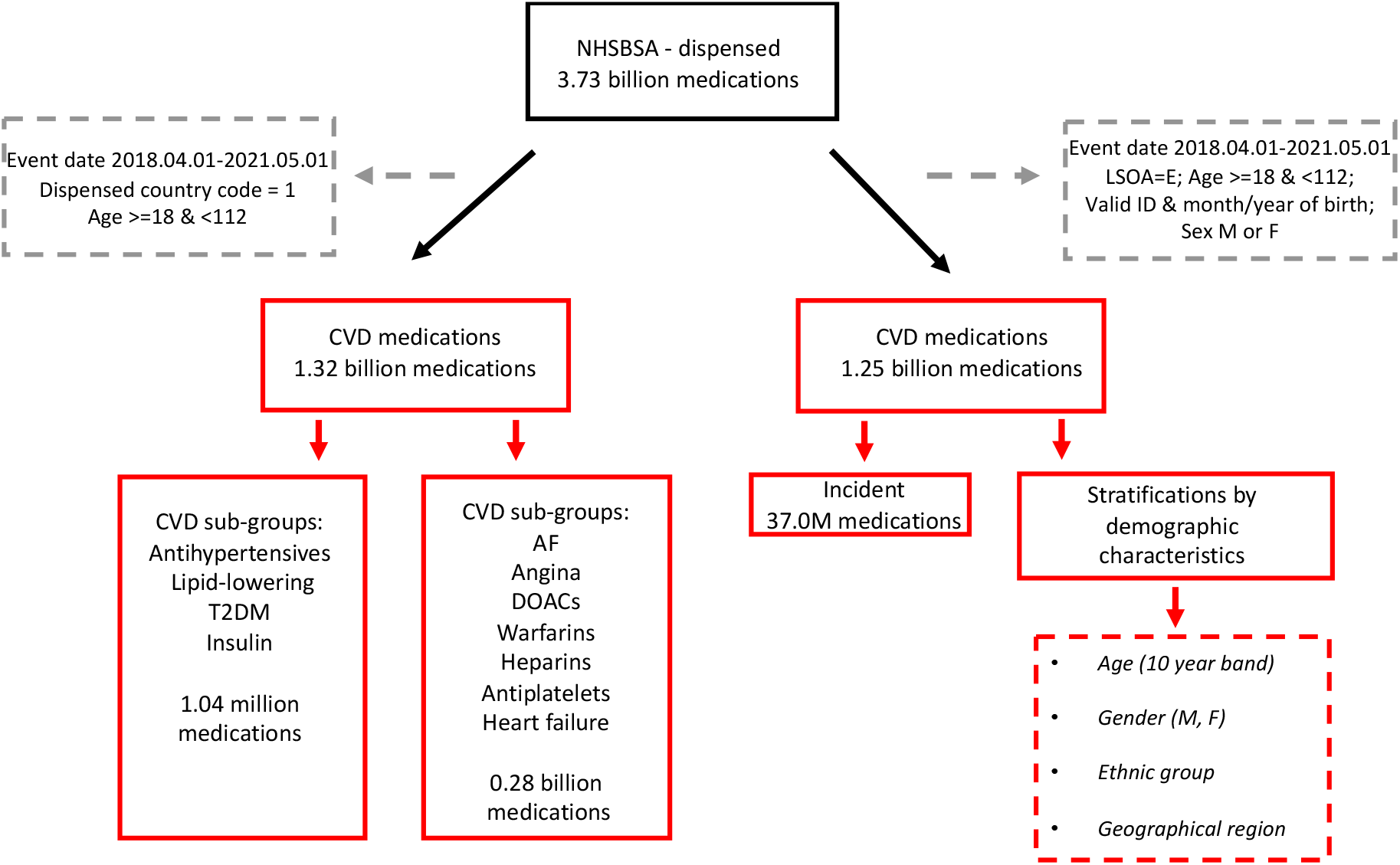
Flowchart showing selection of analytical datasets from NHSBSA (England)

### Study Population

#### Inclusion criteria

The focus of these analyses was medicines with linkage to individual person data for demographic characteristics (**Figure 1**). We included medications dispensed to individuals who were aged between 18 and 112 years, with sex reported as male or female and at pharmacies in the relevant nation (for example, in England Lower-layer Super Output Area (LSOA) starting with E). We excluded individuals with a date of death recorded before 1^st^ April 2018 or a null date of birth. Medications dispensed between 1^st^ April 2018 and 31^st^ July 2021 were included in all three nations. For stratified and incident analyses medication records were required to have a valid pseudo-identifier ID (a non-identifying unique master key that replaces the NHS number following linkage) for data linkage to socio-demographic and regional characteristics. For incident medications, we allowed an initial clearance window for the first year of data availability; incident medications results are therefore presented from 1^st^ March 2019. See **Figure 1** for a flowchart of the selection of data for analysis, linkage, and corresponding counts from data sources.

#### Stratifications

Further stratifications were undertaken according to key demographic characteristics of interest, including by: age (categorised >=18-29 and thereafter in 10 year age bands to 90+ years), sex, ethnicity (categorised as Black, White, Asian, Mixed, Other) and region (categorised as East Midlands, East of England, London, North East, North West, South East, South West, West Midlands, Yorkshire and The Humber, plus Scotland and Wales). Individuals with missing values for a given stratification variable are reported as a separate group for those sub-analyses.

### Statistical Analyses

#### Rates of dispensed medications

We calculated the change in annual rate of dispense for the medicines of interest measuring from 1^st^ March 2018 to end February 2021 (reflecting key timings in the course of the pandemic) for the years 2018-19, 2019-20 and 2020-21. We also calculated year-on-year monthly percentage change in dispensed medications. Results are presented by individual CVD sub-groups and for the combined group of major CVD risk factors: antihypertensives, lipid-lowering medications, T2DM and insulin. Combined counts for the remaining seven CVD sub-groups are available in the **Supplementary Material** (AF, angina, DOACs, warfarins, heparins, antiplatelets and heart failure). Population denominator counts are based on unique individuals receiving at least one dispensed CVD medication in the relevant year (1^st^ March to end February inclusive). Using these denominators, we calculate the annual rate of dispensed medicines per person per year.

#### Stratification by CVD/ CVD risk factor

Monthly counts and year-on-year percent change in monthly counts were calculated for each of the 11 CVD sub-groups for both prevalent and incident medications. We presented results from four of these sub-groups (antihypertensives, lipid-lowering medications, T2DM and insulin) with results for the other seven sub-groups available in the **Supplementary Material** (AF, angina, DOACs, warfarins, heparins, antiplatelets and heart failure).

#### Interrupted time series analyses

To further evaluate trends in medications over time including identification of key periods of change during the pandemic we used Interrupted Time Series (ITS) regression analysis to model the population-level changes in prescribing trends from April 2018 to May 2021, by individual CVD sub-group^18^. Prescription data were required for these analyses to provide greater resolution over time (specifically weekly time points) and hence ITS analysis is restricted to England.

#### Trends in dispensed CVD medications stratified by individual demographic characteristics

For data where it was possible to link medications to individual-level characteristics, we carried out additional analyses to investigate if the variation in dispensing of medications was associated with age, sex, region or ethnicity. A valid NHS ID is required for linkage with individual demographic characteristics; the proportion of data linked improves over time within the English dispensed data (**Supplementary Figure 1**) and this should therefore be considered when interpreting trends by strata.

### Incident CVD medications

We identified the first recorded occurrence of a dispensed medicine from each CVD sub-group within an individual during the study period March 2019 to May 2021. We allowed an initial clearance window for 12 months prior to March 2019 to correct for the high levels of apparent “incidence” in the first few months of data availability. This facilitated identification of within-person incidence at the level of the CVD sub-group; it did not however equate to incidence for the wider CVD medicines group as a whole since individuals may be incident for more than one of the CVD sub-groups.

### Impact of missed treatment on future CVD events

Whilst a full economic analysis was out of scope for this analysis, taking hypertension as an example, we estimated the potential impact of missed cardiovascular risk factor treatment on CVD events using the most recent cost-effectiveness analysis model developed for the National Institute of Health and Care Excellence (NICE) (NICE guideline NG136)^19^, adapting the base case to reflect characteristics of the missed incident hypertensive population. For this we identified characteristics of the 2019 incident population for antihypertensives (mean age, proportion sex, T2DM, smoking) within the English TRE. Using this information in the QRISK2 calculator^20^ we calculated weighted 10-year QRISK2% scores for the NICE treatment effect model base case, additionally specifying SBP at 150mmHg (the threshold for stage 2 antihypertensive treatment using home blood pressure monitoring). Inputting these 10-year QRISK2% scores into the NICE model, we calculated the number of CVD events expected with and without hypertensive treatment (including stratification by stable and unstable angina MI, transient ischaemic attack, stroke and heart failure). We scaled the number of CVD events per 1000 to the missed incident medicines for the treatment of hypertension observed in our analyses across England, Scotland and Wales. We focused on hypertension as the most common CVD risk factor and the most common CVD risk factor for which medicines are prescribed.

## Results

### Overall trends in the dispensing rate of medications

Background rates of CVD medicines dispensed per person per year from March 2020-February 2021 versus the previous year varied by country, increasing in England and Scotland but decreasing in Wales (**Table 1**). In England rates increased from 26.0 to 27.0 CVD medicines per person per year, equating to almost one additional medicine dispensed per person per year (**Table 1)**. This increase can predominantly be accounted for by an increase in the four CVD medicines for hypertension, dysliplidemia and diabetes which increased from 21.4 to 22.4 medicines per person per year in England. Rates for other CVD sub-groups increased less (14.5 to 14.9 medicines per person per year). In contrast, in Wales dispensed rates per person per year fell by a similar amount from 32.8 to 32.0 CVD medicines per person per year, while annual rates in Scotland were more stable increasing by 0.4 to 18.9 medicines per person per year.

**TABLE 1:** Rates and rate differences of CVD medicines dispensed by year (1st March to end February); all CVD medicines and split by CVD sub-group categories; including England, Scotland & Wales.

| Year | ENGLAND |  |  |  | SCOTLAND |  |  |  | WALES |  |  |  |
| --- | --- | --- | --- | --- | --- | --- | --- | --- | --- | --- | --- | --- |
|  | N dispensed items | Patient population | Dispensed medicines rate per person | Year-on-year difference in annual dispensed rate | N dispensed items | Patient population | Dispensed medicines rate per person | Year-on-year difference in annual dispensed rate | N dispensed items | Patient population | Dispensed medicines rate per person | Year-on-year difference in annual dispensed rate |
| *All CVD medicines* |  |  |  |  |  |  |  |  |  |  |  |  |
| March 2018-2019 | 310,280,135 | 13,082,313 |  |  | 26,372,752 | 1,409,313 | 18.7 |  | 26,300,819 | 802,271 | 32.8 |  |
| March 2019-2020 | 351,193,042 | 13,523,877 | 26.0 |  | 26,624,983 | 1,436,342 | 18.5 | -0.2 | 26,810,014 | 817,064 | 32.8 | 0.0 |
| March 2020-2021 | 366,898,585 | 13,573,092 | 27.0 | 1.1 | 26,853,674 | 1,419,592 | 18.9 | 0.4 | 25,875,846 | 808,603 | 32.0 | -0.8 |
| *4 CVD medicines - antihypertensives, lipid-lowering medications, type-2 diabetes, insulin* |  |  |  |  |  |  |  |  |  |  |  |  |
| March 2018-2019 | 243,910,196 | 12,553,549 |  |  | 20,250,786 | 1,333,573 | 15.2 |  | 20,591,866 | 767,947 | 26.8 |  |
| March 2019-2020 | 277,320,496 | 12,980,817 | 21.4 |  | 20,536,503 | 1,358,874 | 15.1 | -0.1 | 21,055,101 | 782,373 | 26.9 | 0.1 |
| March 2020-2021 | 290,942,608 | 13,009,773 | 22.4 | 1.0 | 20,802,410 | 1,343,916 | 15.5 | 0.4 | 20,401,953 | 773,865 | 26.4 | -0.5 |
| *7 CVD medicines - AF, angina, DOACs, warfarins, heparins, antiplatelets, heart failure* |  |  |  |  |  |  |  |  |  |  |  |  |
| March 2018-2019 | 66,369,939 | 5,013,433 |  |  | 6,121,966 | 597,163 | 10.3 |  | 5,708,953 | 325,696 | 17.5 |  |
| March 2019-2020 | 73,872,546 | 5,104,261 | 14.5 |  | 6,088,480 | 597,491 | 10.2 | -0.1 | 5,754,913 | 325,498 | 17.7 | 0.2 |
| March 2020-2021 | 75,955,977 | 5,103,110 | 14.9 | 0.4 | 6,051,264 | 591,229 | 10.2 | 0.0 | 5,473,893 | 320,028 | 17.1 | -0.6 |

English annual patient population is total N unique individuals generated from CVD dataset with valid ID; item counts are therefore also drawn from the same dataset to reflect population at risk No dispensed data are available in English TRE for March 2018; English rates for year 2018-2019 are therefore not directly comparable and excluded from this table

There was an increase in total items of medications dispensed for the common CVD risk factors of hypertension, dysliplidemia and diabetes in the immediate pre-pandemic period (dispense +11.8% March 2020 vs. March 2019) (**Figure 2; Supplementary Table 1**). This compared to year-on-year monthly percentage change ranging between −1.4 and 4.9% in the year before pandemic onset. Year-on-year dispensed percentage did not fall below 2019 levels until May 2020 when initial lockdown restrictions were beginning to be relaxed. Dispensed items again fell below 2019 levels in August 2020 (−9.3%), October 2020 (−1.2%) ahead of the second national lockdown and November 2020 (−0.3%). In comparison, year-on-year dispensing was 4.7% higher in December 2020 ahead of the third national lockdown. The number of medicines was below the previous year throughout early 2021 until April. Overall, we observe a downward trend in the annual percent change in CVD medicines dispensed over the course of 2020 and into 2021 suggesting a decline in the active management of CVD in the population (**Figure 2**).

**FIGURE 2:**
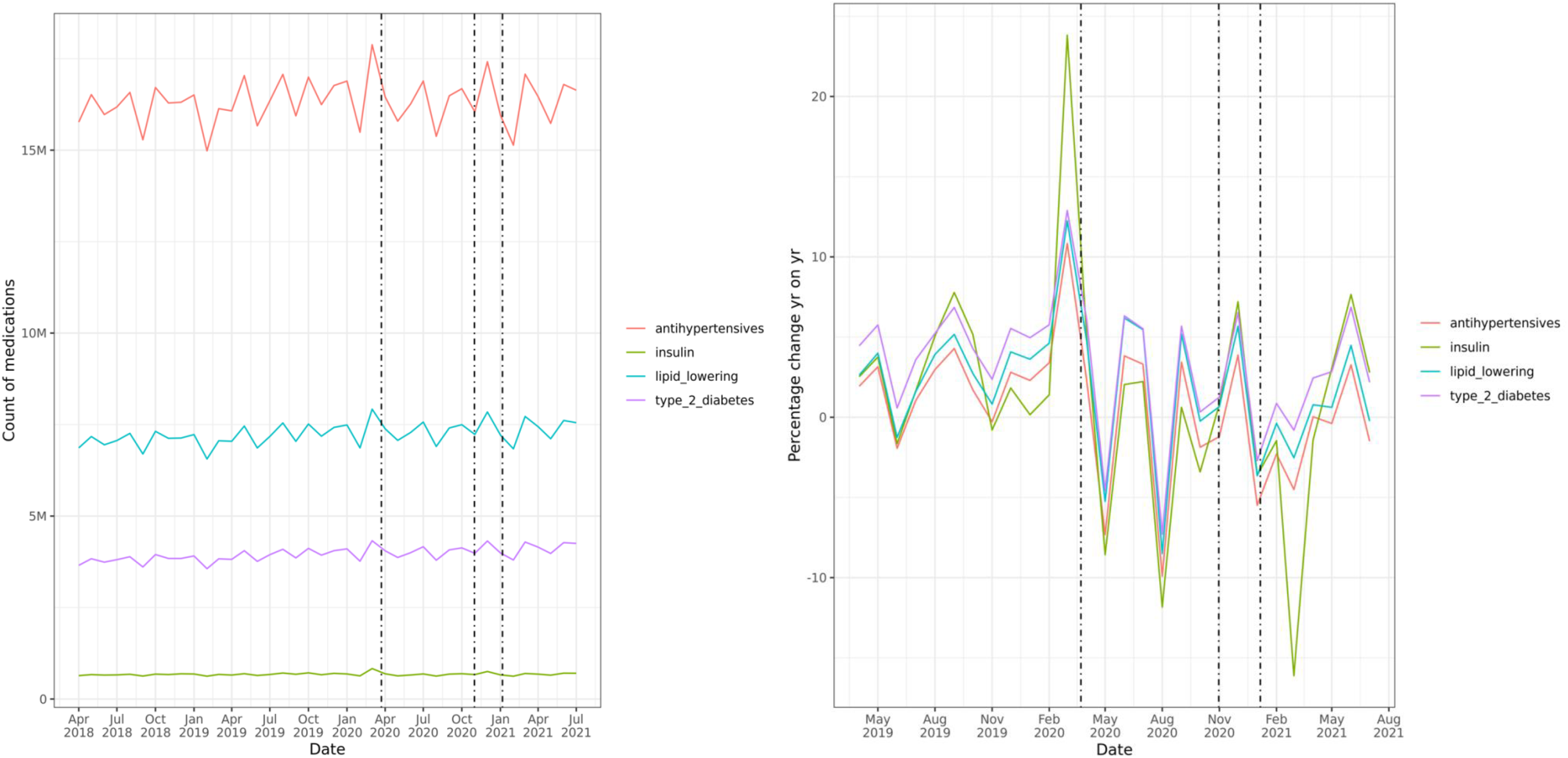
Trends in dispensed CVD medicines over course of pandemic by CVD/ CVD risk factor sub-groups; i) counts by month, and ii) percentage change year-on-year by month; including England, Scotland and Wales. <u>Footnotes</u>: Dotted lines indicate timing of first, second and third national lockdowns: 26^*th*^ March 2020, 5^*th*^ November 2020 and 6^*th*^ January 2021 respectively

### Trends by CVD risk factor, proxied by prevalent medicines

A general pattern of sharp growth in year-on-year medicines dispensed in the pre-pandemic period followed by dispensing below 2019 levels in May 2020 is seen across the CVD sub-groups (**Figure 2**). The most marked year-on-year spike was observed for insulin at +24% in March 2020, followed by dispensing levels below 2019 in May and August 2020. Marked changes were also observed for dispensing of anticoagulant medicines with an acceleration in the decline in warfarin during 2020-21 after an initial spike in March 2020 (**Supplementary Figure 2**). In contrast DOAC dispensing maintained year-on-year growth, but the rate of growth declined (**Supplementary Figure 2**).

### Interrupted time series analyses

We observed a sharp increase in the prescription of CVD medicines in England prior to the first national lockdown, which was similar to increases characteristically observed prior to Christmas (**Figure 3a**). However, unlike Christmas there was no clear subsequent drop in medications prescribed in the week(s) immediately following. The period between the first and second national lockdowns was characterised by declining CVD prescriptions, and, unlike the first lockdown, there was no clear uplift in CVD prescriptions observed in the four-week period preceding the second national lockdown (**Figure 3b**). There was some evidence that the third national lockdown was preceded by a week of uplift; although the period overlap with Christmas and New Year fluctuations hinders interpretation. A similar pattern is observed across all CVD sub-groups.

**FIGURE 3a:**
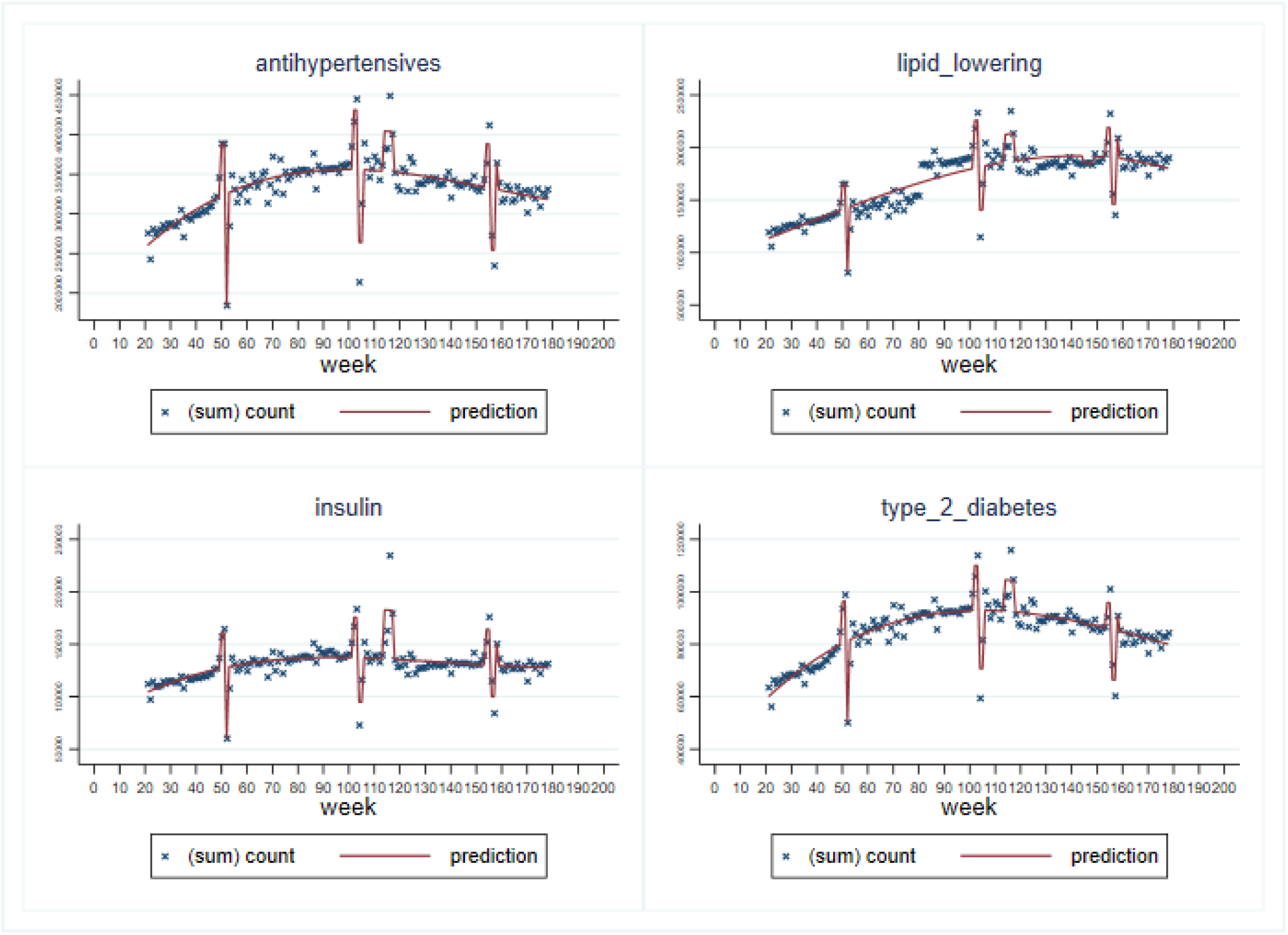
Interrupted time series analysis of prescribed CVD medicines (weekly counts) April 2018 to May 2021, by CVD risk factor; England. <u>Footnotes</u>: Prescribed medicines are analysed for weekly counts; lower counts at weeks 52, 104-5, 154-5 correspond to Christmas & New Year period 2018, 2019, 2020. Peak at weeks 114-117 corresponds to first national lockdown in England. The following sub-periods were classified in the ITS regression: two weeks before Christmas (2018, 2019, 2020), Christmas & New Year period (2018-19, 2019-20, 2020-21), four weeks before national lockdowns (first: 23^*rd*^ March 2020, second: announced 31^*st*^ October 2020 for 5^*th*^ November 2020, third: 6^*th*^ January 2021 – the latter is only one week in duration due to overlap with the Christmas & New Year period 2020-21). Medication data prior to March 2020 and other remaining weeks were used to establish the trend, providing a comparison against which to evaluate the impact of the pandemic. A strength of ITS is that it is typically less affected by common confounding variables (e.g. age, sex etc.) as these change slowly over time. Nevertheless, this method can be affected by time-varying confounders that can change rapidly^*14*^. For this reason, we adjusted for seasonality, as it can be considered a time-dependent confounder in our analysis, using linear, quadratic and cubic functions of time (in weeks). Cubic models provided the best fit for the overall prescribed CVD medicines model and are therefore presented for all CVD sub-groups

**FIGURE 3b:**
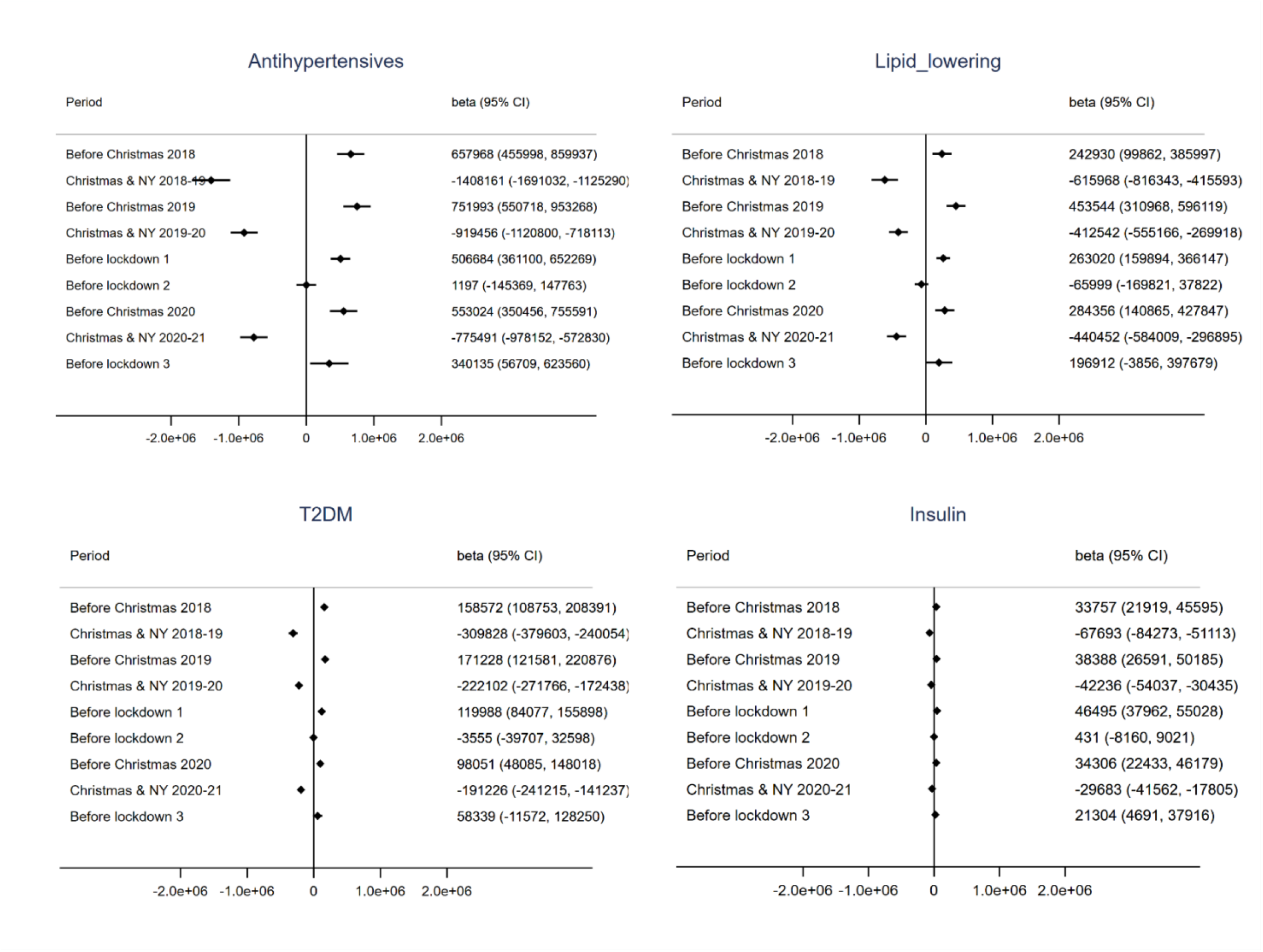
Forest plots of interrupted time series analysis coefficients; prescribed medicines April 2018 to May 2021, by CVD risk factor; England. <u>Footnote</u>: Beta coefficients (& 95% CIs) of change in count of prescribed medicines associated with specified time period. Interrupted time series analysis additionally adjusted for t, t^*2*^ and t^*3*^.

### Dispensing trends by individual demographic characteristics

Data were missing for 6.5% of dispensed CVD medications for region and 1.6% for ethnicity. The highest year-on-year uplifts ahead of the first national lockdown were observed in the age bands 18-29 and 30-39 (**Supplementary Figures 3a & b**). Similar patterns were observed in males and females. Yorkshire and The Humber saw the most pronounced year-on-year uplift in dispensed medicines associated with the first national lockdown and further subsequent peaks in June-July, September reflecting additional local restrictions at those times. London also saw more marked uplifts for subsequent peaks compared to other regions including in December coinciding with the earlier local introduction of Tier 4 restrictions^21^. Similar trends were observed in Scotland and Wales with marked change in year-on-year dispensing associated with the first national lockdown. The Black ethnic group had a delayed uplift in dispensing with year-on-year growth peaking in April 2020 rather than March. A more detailed breakdown by ethnic group is available in England (see **Supplementary Figure 3c**).

### Trends for incident CVD medications

We observe a marked decrease in incident dispensing for antihypertensives, lipid-lowering medications and T2DM medications in the immediate post-pandemic period (**Figure 4**). The easing of lockdown restrictions in May 2020 was followed by a slow recovery in incident prescriptions, but this recovery plateaued with the second and third national lockdowns (5^th^ November 2020 and 6^th^ January 2021 respectively). Incident medications continued to recover through the first half of 2021, with a spike in March 2021 coinciding with the end of the “stay at home” message; however, levels remain markedly lower than in the pre-pandemic period. On average 23,909 fewer patients per month were being commenced on antihypertensives and 14,793 fewer patients on lipid-lowering medications per month during the first half of 2021 compared with the same months in 2019 (**Table 2**). Interestingly, the equivalent change for T2DM is 1,642 more incident patients per month.

**TABLE 2:** Difference in incident medications dispensed by month by CVD/ CVD risk factor sub-group; 2020-21 versus 2019; England, Scotland and Wales.

|  |  | Antihypertensives | Lipid lowering | Type 2 diabetes | Insulin |
| --- | --- | --- | --- | --- | --- |
| March | 2020 | 19,612 | 3,915 | 1,757 | 406 |
| April | 2020 | 44,266 | 29,944 | 10,845 | 1248 |
| May | 2020 | 59,983 | 42,831 | 15,020 | 2,159 |
| June | 2020 | 43,334 | 31,708 | 8,963 | 1028 |
| July | 2020 | 31,900 | 25,733 | 6,094 | 550 |
| August | 2020 | 35,657 | 25,638 | 6,650 | 1,384 |
| September | 2020 | 25,805 | 16,236 | 2,108 | 219 |
| October | 2020 | 29,389 | 17,519 | 2,243 | 355 |
| November | 2020 | 25,070 | 16,411 | 1155 | 15 |
| December | 2020 | 14,857 | 9,807 | -1,044 | -435 |
| January | 2021 | 31,726 | 20,410 | 2,147 | 534 |
| February | 2021 | 20,459 | 18,839 | -691 | -604 |
| March | 2021 | 26,118 | 18,122 | -2,074 | -181 |
| April | 2021 | 22,498 | 14,155 | -2,608 | -229 |
| May | 2021 | 26,394 | 13,520 | -1,225 | 35 |
| June | 2021 | 16261 | 3711 | -5403 | -912 |
| July | 2021 | 17,874 | 5,771 | -3,814 | -353 |
| **Total** |  | **491,203** | **314,270** | **40,123** | **5,219** |

*Differences generated by subtracting 2020-21 monthly incident treatment counts from those observed in 2019 before onset of pandemic*

**TABLE 3:** Estimated N CVD events resulting from missed antihypertensive initiation since March 2020 including data from England, Scotland and Wales; A) assuming non-treatment ongoing over lifetime, and B) non-treatment duration of five years.

| QRISK2% Treatment effect |  |  | Estimated N CVD events in "missed" treatment initiation population |  |  |  |  |  |  |
| --- | --- | --- | --- | --- | --- | --- | --- | --- | --- |
|  |  |  | Stable Angina | Unstable Angina | MI | Transient Ischaemic Attack | Stroke | Heart Failure | Total CVD events |
| A) Lifetime |  |  |  |  |  |  |  |  |  |
| Male | 11.3 | NT | 21,613 | 7,132 | 15,994 | 6,268 | 22,261 | 16,642 | 89,910 |
|  |  | Tx | 19,452 | 6,484 | 14,481 | 5,835 | 21,181 | 16,210 | 83,642 |
|  |  | Additional cases pandemic (NT-Tx) | 2,161 | 648 | 1,513 | 432 | 1,081 | 432 | 6,268 |
| Female | 4.9 | NT | 15,129 | 3,851 | 6,602 | 6,877 | 25,857 | 12,653 | 70,969 |
|  |  | Tx | 13,090 | 3,295 | 5,834 | 6,014 | 23,464 | 11,881 | 63,577 |
|  |  | Additional cases pandemic (NT-Tx) | 2,040 | 556 | 768 | 863 | 2,393 | 772 | 7,392 |
| Total |  |  | 4,201 | 1,205 | 2,281 | 1,295 | 3,474 | 1,204 | 13,659 |
| B) 5 years |  |  |  |  |  |  |  |  |  |
| Male | 11.3 | NT | 3,458 | 1,297 | 3,458 | 648 | 1,513 | 865 | 11,239 |
|  |  | Tx | 3,026 | 1,081 | 2,810 | 648 | 1,297 | 648 | 9,510 |
|  |  | Additional cases pandemic (NT-Tx) | 432 | 216 | 648 | 0 | 216 | 216 | 1,729 |
| Female | 4.9 | NT | 2,000 | 720 | 492 | 985 | 1,409 | 385 | 5,992 |
|  |  | Tx | 1,683 | 606 | 414 | 814 | 1,166 | 322 | 5,005 |
|  |  | Additional cases pandemic (NT-Tx) | 317 | 114 | 78 | 170 | 244 | 63 | 986 |
| Total |  |  | 749 | 330 | 726 | 170 | 460 | 279 | 2,715 |

**FIGURE 4:**
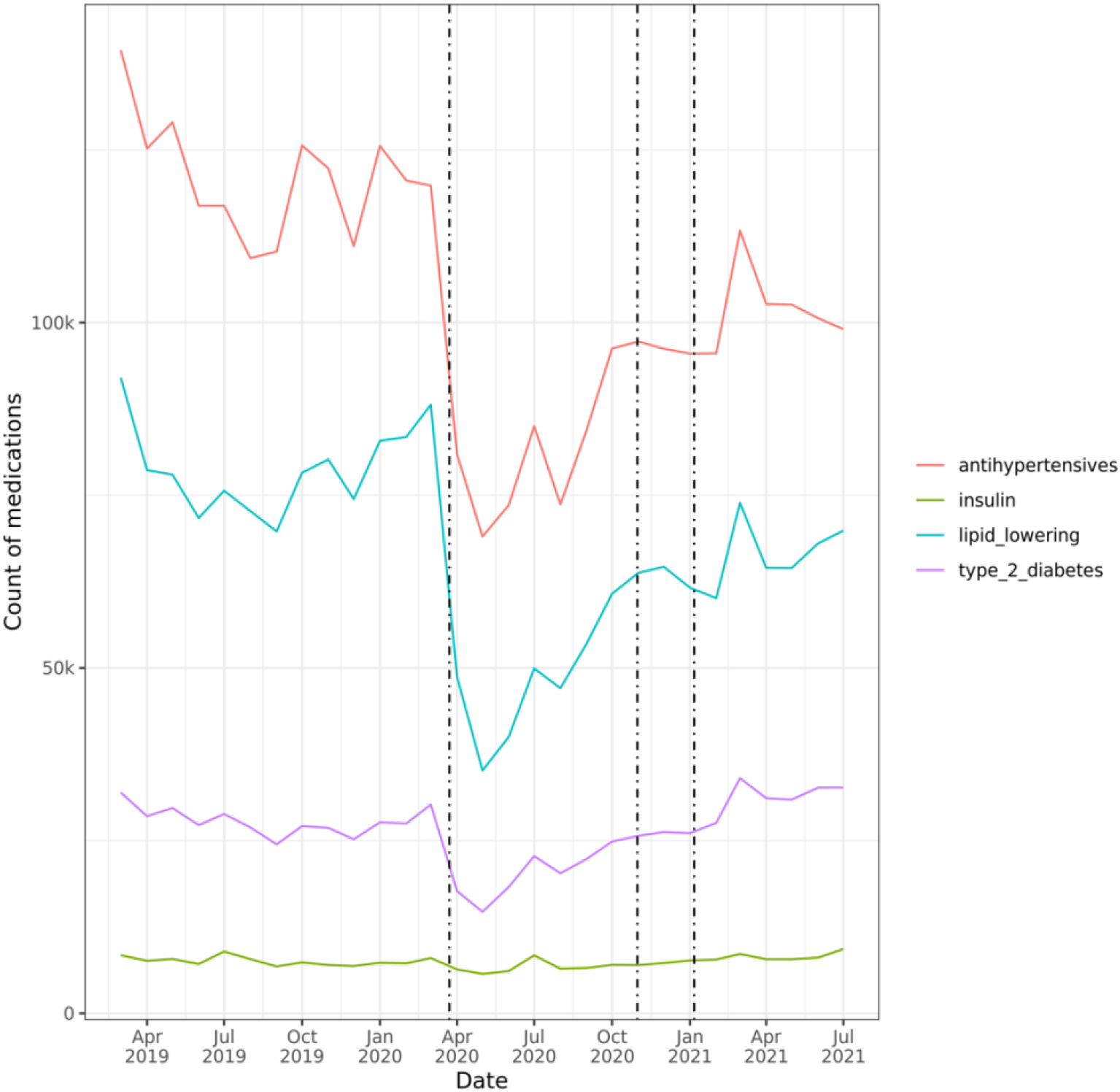
Trends in incident medications dispensed by CVD/ CVD risk factor sub-group; counts by month; England, Scotland and Wales. <u>Footnotes</u>: Dotted lines indicate timing of first, second and third national lockdowns: 26^*th*^ March 2020, 5^*th*^ November 2020 and 6^*th*^ January 2021 respectively.

### Impact of missed treatment on future CVD events

During the period March 2020 to end May 2021 491,203 fewer individuals initiated antihypertensive treatment across England, Scotland and Wales than would have been expected had 2019 incident treatment levels sustained. Using the NICE hypertension treatment model^19^ we estimate that 13,659 additional CVD events will result from the non-initiation of hypertension treatment associated with the COVID-19 pandemic, assuming that this non-treatment were to remain uncorrected for the duration of these individuals’ lives (**Table 4**). This would equate to an additional 2,281 myocardial infarctions and 3,474 strokes resulting from the under-treatment of hypertension alone associated with the COVID-19 pandemic during the period March 2020 to July 2021. Assuming the missed individuals were identified for treatment after five years reduces the total number of CVD events associated with the pandemic to 2,715 CVD events; suggesting that at least 1,555 myocardial infarctions and 3,014 strokes can be avoided if the individuals who have missed incident treatment due to the pandemic can be identified and commenced within this timespan. CVD outcomes for other risk factors (e.g. lipid lowering, T2DM medications) are not estimated here, nor was the additive risk of having one or more of the CVD risk factors which would increase an individual’s absolute risk of a CVD event. In addition, we only consider first treatment with any antihypertensive (rather than specific medicines); this will therefore incorporate individuals started on more than one agent as well as those commenced on monotherapy. As such, this represents conservative estimates of CVD events associated with non-treatment of CVD risk factors due to the pandemic.

## Discussion

### Statement of principal findings

This is the first time that nationwide English, Scottish and Welsh data have been used to describe patterns in dispensed medications. We have used this unique capability to describe how CVD medication has changed during the course of the COVID-19 pandemic. There were marked differences in percentage year-on-year dispensing of CVD medicines by month in 2020-21, including notable uplifts ahead of the first (11.8% higher) but not subsequent national lockdowns. We estimate the number of individuals who are likely to have missed having a major cardiovascular risk factor treated during the course of the COVID-19 pandemic. Compared to pre-pandemic levels in 2019, incident use of antihypertensives in early 2021 fell by an average of 23,909 per month and incident lipid lowering medicines by 14,793 per month. Using hypertension as an example we observed 491,203 fewer individuals initiating antihypertensive treatment across England, Scotland and Wales during the period March 2020 to end May 2021 than would have been expected compared to 2019. An estimate of the additional future impact resulting from this under-treatment of hypertension alone is 13,659 additional cardiovascular events if these individuals remain untreated, equating to 2,281 additional MIs and 3,474 additional strokes. Our data demonstrate that whilst there has been some recovery in dispensing of medications from the initial declines following the first lockdown, crucial first detection of most CVD risk factors as proxied by medicines has not yet returned to pre-pandemic levels. The numbers presented here are likely to be underestimates of the indirect effect of the COVID-19 pandemic as they are based only only on hypertension; a fuller analysis of the impact would need to include all CVD categories.

Use of incident medicines to treat T2DM increased by 1,642 patients per month in the first half of 2021 despite the likely reduced detection, potentially reflecting an increase in new onset T2DM in the population as a result of the events of 2020-21and/ or an awareness of the additional risk of COVID-19 amongst the population and GPs for those with T2DM. This is supported by other evidence pointing to an increase in CVD risk factors within the UK population since the start of the pandemic: for example, BMI is estimated to have increased by ∼+1kg/m^2 22^. Other lifestyle factors not captured through medicines might have increased over the course of the pandemic. Data collected through the ZOE app, suggest that in addition to weight gain, other adverse lifestyle factors reported to have increased include snacking (35%), reduced physical activity (34%) and alcohol consumption (27%) which will further contribute to the risk of hypertension, dyslipidaemia and T2DM^22^. Evidence from other countries also suggests CVD risk factors may have increased during the course of the pandemic, including hypertension^23^. This risk factor increase is not measured in our analyses and therefore may further underestimate the indirect impact of COVID-19 on CVD risk management.

Detection and treatment of CVD risk factors in primary care is usually through routine care of established disease and via vascular health checks and opportunistic screening with mechanisms such as: the Quality of Outcomes Framework (QOF) in England^24^, the Quality Assurance and Improvement Framework (QAIF) in Wales^25^, the Transitional Quality Arrangements (TQA) Framework in Scotland^26^. During the pandemic, early analyses demonstrated that primary care visits fell by 30% from March to June 2020, with many existing visits being replaced by electronic or telephone consultations^2,27^; and the total number of primary care appointments was 10% lower in 2020-21 compared to 2019-2020^3^. This decrease also mirrors a decrease in acute CVD events presenting to secondary care^28^. During different stages of the pandemic there was a re-opening of services, although standard mechanisms of screening risk factors have not been wholly re-introduced^29^. The proportion of face-to-face appointments dropped from 79% to 54%^4^. Declines in consultation rates varied by age, ethnicity and region^3^; with some sub-groups known to have a higher risk of CVD and risk factors associated with CVD^30^, including men, less affluent patients and immigrants, are less likely to access remote consultations^31^. The decline in treatment of incident CVDs we observe might be explained by suspension of incentivised screening and routine reviews such as pay for performance (QOF) as well as a reduction in face-to-face consultations as many appointments continue to be remote^27^.

### Strengths and weaknesses of the study

This is the largest ever study to date using a medicines lens to investigate the impact of the COVID-19 pandemic on CVD risk factors at a population-scale in England, Scotland and Wales. A strength of using a medicines perspective is that it is closer to the active control of CVD risk factors in the patient population. We developed and applied a new categorisation of CVD medicines according to prescribed medicine use for this purpose informed by pharmacological and clinical knowledge of use of these medicines in practice. A limitation relates to the difficulty in assigning diseases for overlapping indications for some medications. The approach we used aligns with the primary indication for a medication as per the BNF. This may result in underestimates of certain CVDs. For example, heart failure is likely to be underestimated as management options overlap with hypertension and type-2 diabetes (e.g. ACE-I, beta blockers and SGLT-2 inhibitors). This analysis could be extended in future work by linking to disease diganosis codes to refine estimates for heart failure. However, the analyses presented here do give an indication of the overall missed CVD risk factors to alert policy makers to the indirect effect of COVID-19.

These analyses do not take account of dispensed quantity with each dispense counting as a single unit and this issue lends itself to further research. We used atorvastatin 40mg and simvastatin 40mg to demonstrate that this is unlikely to be affecting these analyses (**Supplementary Table 5**), as quantities of these medicines remained stable over the course of the pandemic. Observed differences in quantity by country may explain variation observed in inter-country dispensing rates. Medication data analysed here represent real world data that were not collected for research purposes; it is possible that artefact may exist within the data due to differences in collection, processing or transfer and these may vary over time and by source. We observed a decline in the proportion of medications dispensed with invalid (“null”) IDs over time in the English data corresponding with an ongoing switch away from paper-based to electronic processing^32^ (**Supplementary Figure 1**); valid IDs are required for linkage with other data including individual characteristics.

### Strengths and weaknesses in relation to other studies, discussing important differences in results

This work complements and meaningfully extends other evidence. An early report from the Health Foundation using data from CPRD indicated that there was an increase prior to the first lockdown of prescribed and dispensed medicines^2^. This is similar to patterns that are often seen, and again demonstrated in these analyses, of accumulation of medicines prior to Christmas to account for the holiday closures. However, unlike Christmas there was no subsequent drop in medications prescribed in the week(s) immediately following. Patients therefore accessed additional medications but continued to have medications prescribed at a similar level to pre-pandemic levels, suggestive of some stockpiling of medications. Welsh data using all community dispensing data found similar patterns for cardiovascular categories until September 2020^16^; data which are also now incorporated into these analyses and updated until Summer 2021. As lockdown eased, a return of some services to near normal levels has been described but this did not include medication data^33^. Lower levels of diagnosis of T2DM during 2020 following the first lockdown in April have previously been reported^4^; the subsequent higher level of incident T2DM observed in these analyses in 2021, despite the known reduction in primary care screening, could suggest an increase in the prevalence of T2DM in the UK population, or that there is now a ‘catch up’ in diabetes diagnosis, which if true means more people are being diagnosed later with more advanced disease. Data from the Office for National Statistics show that deaths were ∼20% higher than the pre-pandemic average in England & Wales during the Summer of 2021 and half of these excess deaths did not directly involve COVID-19^34^, suggesting that other causes of death, including CVD, are higher than they were pre-pandemic.

Since COVID-19 has highlighted a number of inequities in healthcare we investigated differences in dispensing behaviour by socio-demographic characteristics. Previous research has demonstrated that individuals of certain ethnic groups may be more at risk of COVID-19^35^ and additionally that CVD conditions can be a major risk factor for COVID-19^36^. If CVD risk factors are systematically being missed and not controlled in higher risk populations (defined by region, ethnicity or other characteristics) this may put these populations at greater risk of future health problems. It is known that there is ethnic variation in prescribing of some CVD risk factor medications. For example, recent analyses using CPRD revealed an ethnic difference in receiving statins, with those of African/ African

Caribbean and South Asian ethnicity less likely to receive statins than those of European ethnicity^37^. It may be that the delayed uplift in dispensed medication associated with the first national lockdown in some black ethnic groups are further manifestation of inequalities in receiving medications. We also observed regional variation in the intensity of fluctuation of dispensing of CVD medicines reflecting the timing of local lockdowns which may be a proxy for deprivation and poorer health. A more granular stratification of sub-regions e.g., ward level within broader regional categories, or by index of multiple deprivation, may allow a better understanding of these differences.

### Meaning of the study: possible explanations and implications for clinicians and policymakers

Our analyses demonstrate that dispensed medicines for some incident CVD risk factors did not recover and still have not recovered to pre-pandemic levels. This further aligns with data demonstrating the fall in GP visits, which precludes serendipitous detection, but also the suspension of mechanisms that serve to detect risk factors such as QOF. Whilst it is likely that as services return to normal, cardiovascular risk in missed individuals may well be detected, it remains unclear what mechanisms are in place to re-introduce methods of screening or what the consequences a delay in diagnosis might have. An important consideration from these analyses is implications more generally about health service provision during pandemics and planning for how routine health care could be sustained despite demands on the overall system in the event of future pandemics. The COVID-19 booster campaign whilst essential has focussed attention away from long term conditions. These analyses may provide mechanisms to identify and then target those at highest CVD risk. However, we must also identify alternative mechanisms of risk factor management incopororating support services in primary care e.g. primary care pharmacists and local pharmacies which may be able to address less complex cases.

Different patterns in certain medicine classes are also revealed. Insulin dispensing rose sharply prior to the first lockdown; it is plausible that individuals requiring insulin would be concerned about potential shortage of the drug during lockdown and therefore stockpiled this to ensure an adequate supply given the immediate adverse effects of non-treatment. The uptake of DOACs was increasing pre-pandemic and the pandemic may have accelerated this uptake and the switch away from warfarin as described elsewhere^33,38^. However, declining year-on-year growth in DOAC dispensing during the pandemic may indicate reduced diagnoses of AF and thrombo-embolic disease. In addition, the reduced level of warfarin may reflect missed diagnoses requiring anti-coagulation with warfarin such as valvular heart disease.

### Unanswered questions and future research

There are many further opportunities to explore with medications data that are beyond the scope of this paper. It is now possible to link dispensing data with GPES at an individual level facilitating detailed analysis of characteristics associated with life-course use and accumulation of medications (polypharmacy), adverse drug reactions and adherence. Further research is warranted to understand the indirect cost of the impact of the pandemic on CVD risk factors and longer-term outcomes. Understanding how medicines are being used can act as a barometer for the ‘health’ or disruption to a clinical pathway and as these analyses demonstrate may also help target recovery. This approach is useful in routine health care outside of a pandemic in addressing medicines use more broadly and assuring equity of access. Work in this area will be advanced at the UK-wide level under a recently awarded HDR UK Alan Turing Institute COVID-19 Data and Connectivity National Core Study grant.

It is clear from these analyses that using medicines as a proxy for disease can complement investigation using electronic health records and disease diagnostic codes. During the pandemic due to the change in health care service provision and reduced use of routine services, a proportion of the expected diagnostic codes are missing, or not clearly coded, including long Covid. Medicines data can therefore be another way to begin to establish the scale of the health problem resulting from the pandemic including beginning to ascertain the cost of treatments that were not given. Novel methods to identify and treat individuals who have missed treatment are urgently required to avoid large numbers of additional future CVD events, further adding to the healthcare impact and indirect cost of the COVID-19 pandemic

## Data Availability

Data used in this study are available in NHS Digitals Trusted Research Environment (TRE) for England, but as restrictions apply they are not publicly available (https://digital.nhs.uk/coronavirus/coronavirus-data-services-updates/trusted-research-environment-service-for-england). The CVD-COVID-UK/COVID-IMPACT programme led by the BHF Data Science Centre (https://www.hdruk.ac.uk/helping-with-health-data/bhf-data-science-centre/) in partnership with HDR UK received approval to access data in NHS Digitals TRE for England from the Independent Group Advising on the Release of Data (IGARD) (https://digital.nhs.uk/about-nhs-digital/corporate-information-and-documents/independent-group-advising-on-the-release-of-data) via an application made in the Data Access Request Service (DARS) Online system (ref. DARS-NIC-381078-Y9C5K) (https://digital.nhs.uk/services/data-access-request-service-dars/dars-products-and-services). The CVD-COVID-UK/COVID-IMPACT Approvals & Oversight Board (https://www.hdruk.ac.uk/projects/cvd-covid-uk-project/)) subsequently granted approval to this project to access the data within the TRE for England, the Scottish National Safe Haven and the Secure Anonymised Information Linkage (SAIL) Databank. The de-identified data used in this study was made available to accredited researchers only.
Data used in this study are available in the Scottish National Safe Haven (Project Number: 2021-0102), but as restrictions apply they are not publicly available. Access to data may be granted on application to the Public Benefit and Privacy Panel for Health and Social Care (PBPP (https://www.informationgovernance.scot.nhs.uk/pbpphsc/)). Applications are co-ordinated by eDRIS (electronic Data Research and Innovation Service (https://www.isdscotland.org/Products-and-services/Edris/)). The anonymised data used in this study was made available to accredited researchers only through the Public Health Scotland (PHS) eDRIS User Agreement (https://www.isdscotland.org/Products-and-services/Edris/_docs/eDRIS-User-Agreement-v16.pdf).
Data used in this study are available in the SAIL Databank at Swansea University, Swansea, UK, but as restrictions apply they are not publicly available. All proposals to use SAIL data are subject to review by an independent Information Governance Review Panel (IGRP). Before any data can be accessed, approval must be given by the IGRP. The IGRP gives careful consideration to each project to ensure proper and appropriate use of SAIL data. When access has been granted, it is gained through a privacy protecting data safe haven and remote access system referred to as the SAIL Gateway. SAIL has established an application process to be followed by anyone who would like to access data via SAIL at https://www.saildatabank.com/application-process.

https://github.com/BHFDSC/CCU014_01

## Public and Patient Involvement

The project was approved by the BHF DSC Approvals & Oversight Board which included public partners, who were also consulted as data were produced and inputted into the final manuscript.

## Ethical approval

The North East-Newcastle and North Tyneside 2 research ethics committee provided ethical approval for the CVD-COVID-UK research programme (REC No 20/NE/0161).

## Software and code availability and data sharing

All data preparation and analyses were conducted using Databricks (SQL, Python), R or Stata within the English TRE. All data preparation and analyses within the Scottish National Safe Haven were conducted on the secure analytical platform using R. All data processing in the SAIL Databank was performed using R. All code is available on GitHub https://github.com/BHFDSC/CCU014_01.

Data used in this study are available in NHS Digital’s Trusted Research Environment (TRE) for England, but as restrictions apply they are not publicly available (https://digital.nhs.uk/coronavirus/coronavirus-data-services-updates/trusted-research-environment-service-for-england). The CVD-COVID-UK/COVID-IMPACT programme led by the BHF Data Science Centre (https://www.hdruk.ac.uk/helping-with-health-data/bhf-data-science-centre/) in partnership with HDR UK received approval to access data in NHS Digital’s TRE for England from the Independent Group Advising on the Release of Data (IGARD) (https://digital.nhs.uk/about-nhs-digital/corporate-information-and-documents/independent-group-advising-on-the-release-of-data) via an application made in the Data Access Request Service (DARS) Online system (ref. DARS-NIC-381078-Y9C5K) (https://digital.nhs.uk/services/data-access-request-service-dars/dars-products-and-services). The CVD-COVID-UK/COVID-IMPACT Approvals & Oversight Board (https://www.hdruk.ac.uk/projects/cvd-covid-uk-project/)) subsequently granted approval to this project to access the data within the TRE for England, the Scottish National Safe Haven and the Secure Anonymised Information Linkage (SAIL) Databank. The de-identified data used in this study was made available to accredited researchers only.

Data used in this study are available in the Scottish National Safe Haven (Project Number: 2021-0102), but as restrictions apply they are not publicly available. Access to data may be granted on application to the Public Benefit and Privacy Panel for Health and Social Care (PBPP (https://www.informationgovernance.scot.nhs.uk/pbpphsc/)). Applications are co-ordinated by eDRIS (electronic Data Research and Innovation Service (https://www.isdscotland.org/Products-and-services/Edris/)). The anonymised data used in this study was made available to accredited researchers only through the Public Health Scotland (PHS) eDRIS User Agreement (https://www.isdscotland.org/Products-and-services/Edris/_docs/eDRIS-User-Agreement-v16.pdf).

Data used in this study are available in the SAIL Databank at Swansea University, Swansea, UK, but as restrictions apply they are not publicly available. All proposals to use SAIL data are subject to review by an independent Information Governance Review Panel (IGRP). Before any data can be accessed, approval must be given by the IGRP. The IGRP gives careful consideration to each project to ensure proper and appropriate use of SAIL data. When access has been granted, it is gained through a privacy protecting data safe haven and remote access system referred to as the SAIL Gateway. SAIL has established an application process to be followed by anyone who would like to access data via SAIL at https://www.saildatabank.com/application-process.

## Acknowledgement

This work is carried out with the support of the BHF Data Science Centre led by HDR UK (BHF Grant no. SP/19/3/34678) and makes use of de-identified data held in NHS Digital’s Trusted Research Environment for England, the SAIL Databank and the Scottish National Data Safe Haven, made available via the HDR UK BHF Data Science Centre’s CVD-COVID-UK consortium. This work uses data provided by patients and collected by the NHS as part of their care and support. We would also like to acknowledge all data providers who make health relevant data available for research.

The study makes use of anonymised data held in the Scottish National Safe Haven. The authors would like to acknowledge the support of the eDRIS Team (Public Health Scotland) for their involvement in obtaining approvals, provisioning and linking data and the use of the secure analytical platform within the National Safe Haven.

This study makes use of anonymised data held in the Secure Anonymised Information Linkage (SAIL) Databank. This work uses data provided by patients and collected by the NHS as part of their care and support. We would also like to acknowledge all data providers who make anonymised data available for research. We wish to acknowledge the collaborative partnership that enabled acquisition and access to the de-identified data, which led to this output. The collaboration was led by the Swansea University HDR UK team under the direction of the Welsh Government Technical Advisory Cell (TAC) and includes the following groups and organisations: the SAIL Databank, Administrative Data Research (ADR) Wales, Digital Health and Care Wales (DHCW), Public Health Wales, NHS Shared Services Partnership (NWSSP) and the Welsh Ambulance Service Trust (WAST). All research conducted has been completed under the permission and approval of the SAIL independent Information Governance Review Panel (IGRP) project number 0911.

## SUPPLEMENTARY METHODS

### Details of Medications Data Processing by nation

#### England

NHSBSA data comprise prescriptions for medicines that are dispensed or supplied by community pharmacists, appliance contractors and dispensing doctors in England. The data also includes prescriptions submitted by prescribing doctors, for medicines personally administered in England.

NHSBSA data were screened to produce a master list of all medications with any record of being dispensed^8^. Medicines are identifiable by a BNF or DM+D code (a dictionary of descriptions and codes which represent medicines and devices of use across the NHS). Medicines were selected on the basis of a unique combination of British National Formulary (BNF) and Dictionary of Medicines and Devices (DM+D) codes. This combination permitted identification of all unique medicines and facilitated mapping to CVD categorisation. Twenty-four apparently illogical duplicate medicines were identified with two different BNF codes per DM+D code; of these we retained only the BNF code with corresponding product information. None of the 24 were identified as CVD medicines and are therefore filtered out as part of our analysis pipeline. 32,574 unique medicines with distinct BNF codes (each specific combination of substance-pack-concentration) were taken forward for analysis. CVD medicines were selected from these using the categorisation described above.

Four hundred and fifteen medications appearing in the possible list of all medications screened from NHSBSA were subsequently identified in GDPPR via their DM+D code. Since medicines are identified by only a single DM+D code in the GDPPR data, but by multiple DM+D codes in the NHSBSA data it was necessary to re-map the selection of these medicines via the relevant unique BNF-DM+D combination and use the corresponding BNF code to select the 415 from the NHSBSA. This selection of 415 medicines is used for direct comparison of trends in prescription and dispense of medicines; while analyses of NHSBSA alone include additional CVD medicines not available in GDPPR.

Dates in NHSBSA reflect the month in which the script was submitted for payment rather than the date a medication was dispensed to the patient; whereas the date variable in the GDPPR data reflects the actual day on which a medication is prescribed by the GP. The first available month of NHSBSA data is April 2018. Prescriptions can be identified in GDPPR as early as 2015 but data before mid-2018 are sporadic and less reliable; we therefore applied an April 2018 start date to the majority of analyses (with the exception of some comparisons where early artefact issues in the early GDPPR data necessitated a later start date of September 2018). The analysis end date was the latest available monthly download at time of analysis for NHSBSA and the most recent prescriptions available in GDPPR at the time of analysis (31^st^ May 2021).

Ages were calculated at the date of dispensing for each medication by subtracting the month and year of birth from the dispense date. For stratified analyses, demographic and regional data were linked from NHS Digital source “skinny” tables via the unique NHS person ID available in the TRE (a unique 10 digit identifier assigned to patients upon first interaction with the healthcare system).

#### Scotland

The Scottish Prescribing Information System (PIS) provides a repository for all community prescribing related information, including payments, but excluding prescriptions dispensed in hospitals.^39^ PIS comprises three different records/sources of data: 1) ePrescibed – details submitted through the prescribing system (usually a GP practice), 2) eDispensed – details submitted through the dispensing system (community pharmacy), 3) DCVP – details used for payment to the pharmacy. The dispensed data in this study contains those prescriptions which have been processed completely through the system from prescription to payment. Prescriptions may be missing from the available dataset if they have not been presented to a community pharmacy for dispensing, if the prescription was not paid for through the NHS (e.g. if it was a private prescription), or if the CHI number was missing for any reason and the record could not be linked to an individual. The dates in the individual records include the date the prescription was issued, the date it was dispensed, and the date payment was made. Dispensed dates are not necessarily real dates but could be default dates, for example the last day of a month. Data are available from April 2009, but the data requested for CVD-COVID-UK projects and available on the Safe Haven is from 1^st^ January 2015 onwards.

Ethnicity is not available as part of the PIS data on the Scottish National Safe Haven. More generally ethnicity has historically not been reliably recorded in Scottish health care records, with the coding scheme used in the Scottish Morbidity Records changing to use Scottish Census 2011 Ethnicity Categories in 2011 (mandatory since 1^st^ April 2012). A different coding scheme is in use by the National Records of Scotland. There have been recent improvements in data collection in response to the UK Government’s Equality Act. This should allow a more accurate categorisation going forward.^40^ Ethnicity data from Scotland are therefore not included in these analyses.

NHS Scotland is currently divided into 14 regional Health boards, with each health board having a large degree of autonomy with regard to how they implement and deliver health services^41^. The geographical size and populations of each region may differ substantially. For example, as of 2019, NHS Greater Glasgow and Clyde serves approximately 1.2 million people, whereas NHS Shetland covers a population of approximately 23,000.^42,43^. Each PIS dispensed record for a patient includes region information via a health board of residence field. In the vast majority of cases this is one of the 14 health boards covering Scotland, but a small number of special codes exist (e.g. No Fixed Abode), covering a very small number of patients who do not have a health board of residence.

#### Wales

Dispensing data: The available range of Welsh Dispensing DataSet (WDDS) at the time of this study was from 1^st^ January 2016 to 25^th^ August 2021. The raw data arrives in two separate extracts, one including all dispensed items per practice (each person within a general practice setting is identified by a unique ID in the data extract) and the other including an anonymised linkage field (ALF) that enables linkage of dispensing records to other available patient information^15^. Within WDDS, all medications are coded in DM+D. We established a pipeline that is applied to each monthly release of WDDS data that links both ALF and Dispensing record tables and maps drug items from DM+D codes to BNF. NHSBSA was used to map all dispensed items from DM+D codes to BNF coding system^44^. Details of mapping strategy and syntax is provided at https://github.com/BHFDSC/CCU014_01. In order to match the existing data range available in England and Scotland, a snapshot of Wales data starting from March-2018 up to May-2021 was used for these analyses.

Prescribing data: The available range of Welsh Longitudinal General Practice (WLGP) data at the time of this study was from 1^st^ January 2000 to 1^st^ October 2021. Although prescribing data exists for Wales population, a detailed clinical review and validation of mapping from Read to BNF are required before being able to make a cross country comparison across all groupings of the prescribed medication in this analysis. We hope to complete this mapping in due course.

Ages were calculated at the date of dispensing for each medication by subtracting the week of birth (Monday) from the dispense date.

Ethnicity data was extracted from a combination of electronic health record data sources, and harmonised into a composite national ethnicity spine which corresponds to the Office for National Statistics (ONS) breakdowns of ethnicity categories for the population of Wales, in the following five groupings: White, Asian, Black, Mixed and Other.

Using a row sequence counting total number of each medication per individual, incident medications were then defined as first appearance of a dispensed medication in a certain time period which for this study was March 2019 onward.

## SUPPLEMENTARY TABLES

**SUPPLEMENTARY TABLE 1:**
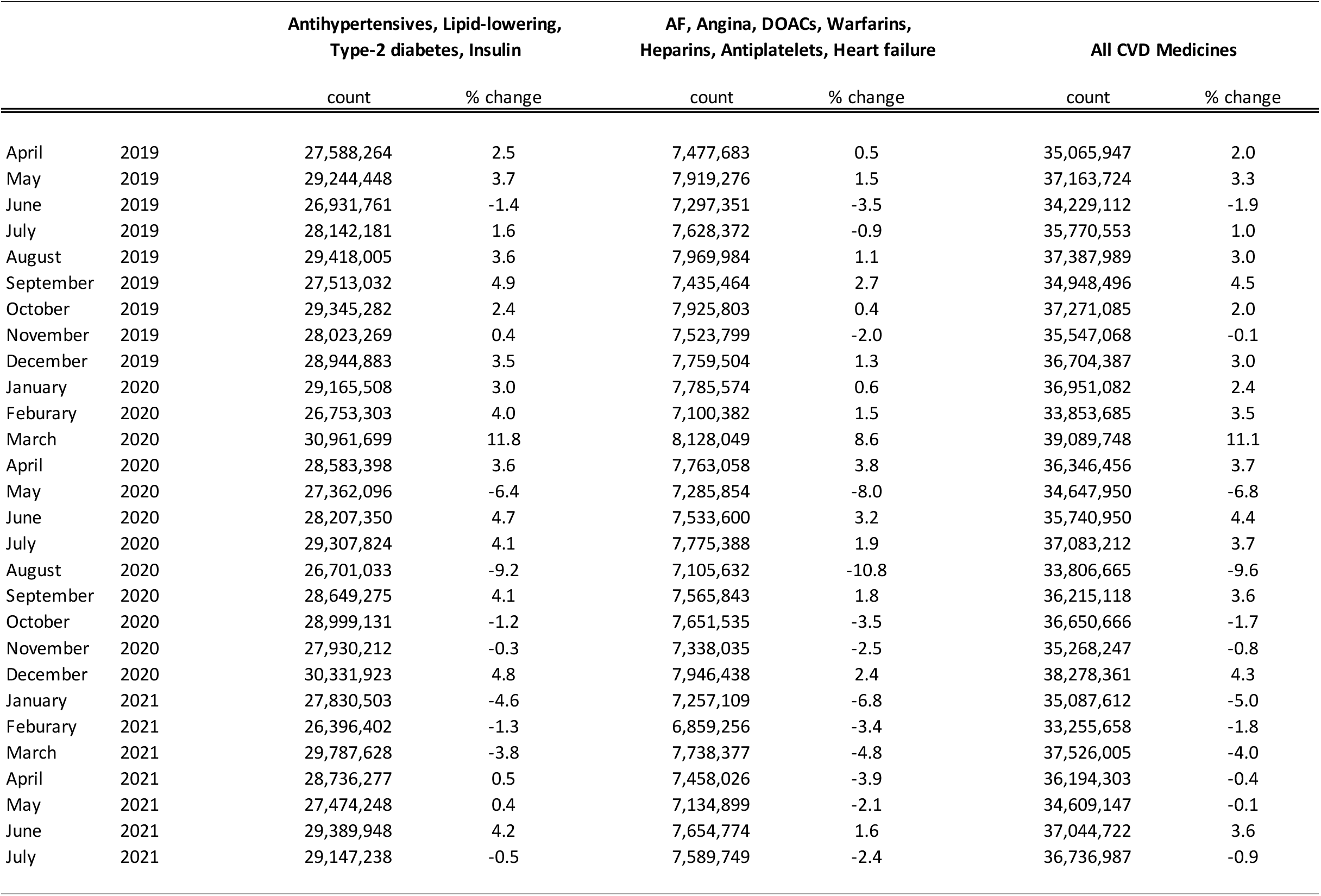
Year-on-year percentage change in monthly counts of CVD medicines; England, Scotland and Wales.

|  |  | Antihypertensives, Lipid-lowering,<br>Type-2 diabetes, Insulin |  | AF, Angina, DOACs, Warfarins,<br>Heparins, Antiplatelets, Heart failure |  | All CVD Medicines |  |
| --- | --- | --- | --- | --- | --- | --- | --- |
|  |  | count | % change | count | % change | count | % change |
| April | 2019 | 27,588,264 | 2.5 | 7,477,683 | 0.5 | 35,065,947 | 2.0 |
| May | 2019 | 29,244,448 | 3.7 | 7,919,276 | 1.5 | 37,163,724 | 3.3 |
| June | 2019 | 26,931,761 | -1.4 | 7,297,351 | -3.5 | 34,229,112 | -1.9 |
| July | 2019 | 28,142,181 | 1.6 | 7,628,372 | -0.9 | 35,770,553 | 1.0 |
| August | 2019 | 29,418,005 | 3.6 | 7,969,984 | 1.1 | 37,387,989 | 3.0 |
| September | 2019 | 27,513,032 | 4.9 | 7,435,464 | 2.7 | 34,948,496 | 4.5 |
| October | 2019 | 29,345,282 | 2.4 | 7,925,803 | 0.4 | 37,271,085 | 2.0 |
| November | 2019 | 28,023,269 | 0.4 | 7,523,799 | -2.0 | 35,547,068 | -0.1 |
| December | 2019 | 28,944,883 | 3.5 | 7,759,504 | 1.3 | 36,704,387 | 3.0 |
| January | 2020 | 29,165,508 | 3.0 | 7,785,574 | 0.6 | 36,951,082 | 2.4 |
| February | 2020 | 26,753,303 | 4.0 | 7,100,382 | 1.5 | 33,853,685 | 3.5 |
| March | 2020 | 30,961,699 | 11.8 | 8,128,049 | 8.6 | 39,089,748 | 11.1 |
| April | 2020 | 28,583,398 | 3.6 | 7,763,058 | 3.8 | 36,346,456 | 3.7 |
| May | 2020 | 27,362,096 | -6.4 | 7,285,854 | -8.0 | 34,647,950 | -6.8 |
| June | 2020 | 28,207,350 | 4.7 | 7,533,600 | 3.2 | 35,740,950 | 4.4 |
| July | 2020 | 29,307,824 | 4.1 | 7,775,388 | 1.9 | 37,083,212 | 3.7 |
| August | 2020 | 26,701,033 | -9.2 | 7,105,632 | -10.8 | 33,806,665 | -9.6 |
| September | 2020 | 28,649,275 | 4.1 | 7,565,843 | 1.8 | 36,215,118 | 3.6 |
| October | 2020 | 28,999,131 | -1.2 | 7,651,535 | -3.5 | 36,650,666 | -1.7 |
| November | 2020 | 27,930,212 | -0.3 | 7,338,035 | -2.5 | 35,268,247 | -0.8 |
| December | 2020 | 30,331,923 | 4.8 | 7,946,438 | 2.4 | 38,278,361 | 4.3 |
| January | 2021 | 27,830,503 | -4.6 | 7,257,109 | -6.8 | 35,087,612 | -5.0 |
| February | 2021 | 26,396,402 | -1.3 | 6,859,256 | -3.4 | 33,255,658 | -1.8 |
| March | 2021 | 29,787,628 | -3.8 | 7,738,377 | -4.8 | 37,526,005 | -4.0 |
| April | 2021 | 28,736,277 | 0.5 | 7,458,026 | -3.9 | 36,194,303 | -0.4 |
| May | 2021 | 27,474,248 | 0.4 | 7,134,899 | -2.1 | 34,609,147 | -0.1 |
| June | 2021 | 29,389,948 | 4.2 | 7,654,774 | 1.6 | 37,044,722 | 3.6 |
| July | 2021 | 29,147,238 | -0.5 | 7,589,749 | -2.4 | 36,736,987 | -0.9 |

**SUPPLEMENTARY TABLE 2:** Rates and rate differences of CVD medicines prescribed by year (1st March to end February); all CVD medicines and split by key CVD sub-groups, England.

| Year | Number of<br>prescribed items | Patient<br>population | Prescribed<br>medicines rate<br>per person | Year on year<br>difference in<br>annual<br>prescribed rate |
| --- | --- | --- | --- | --- |
| *England* |  |  |  |  |
| *All CVD medicines* |  |  |  |  |
| March 2018-2019* | 258,190,285 | 11,920,436 |  |  |
| March 2019-2020 | 401,092,906 | 12,946,298 | 31.0 |  |
| March 2020-2021 | 400,819,210 | 13,049,342 | 30.7 | -0.3 |
| *4 CVD medicines - antihypertensives, lipid-lowering medications, type-2 diabetes, insulin* |  |  |  |  |
| March 2018-2019* | 212,386,072 | 11,507,573 |  |  |
| March 2019-2020 | 329,979,852 | 12,474,219 | 26.5 |  |
| March 2020-2021 | 329,573,385 | 12,536,678 | 26.3 | -0.2 |
| *7 CVD medicines - AF, angina, DOACs, warfarins, heparins, antiplatelets, heart failure* |  |  |  |  |
| March 2018-2019* | 45,804,213 | 4,249,494 |  |  |
| March 2019-2020 | 71,113,054 | 4,672,654 | 15.2 |  |
| March 2020-2021 | 71,245,825 | 4,787,037 | 14.9 | -0.3 |

No GPPR data are available in English TRE for March 2018; rates for year 2018-2019 are therefore not directly comparable and excluded from this table

**SUPPLEMENTARY TABLE 3:**
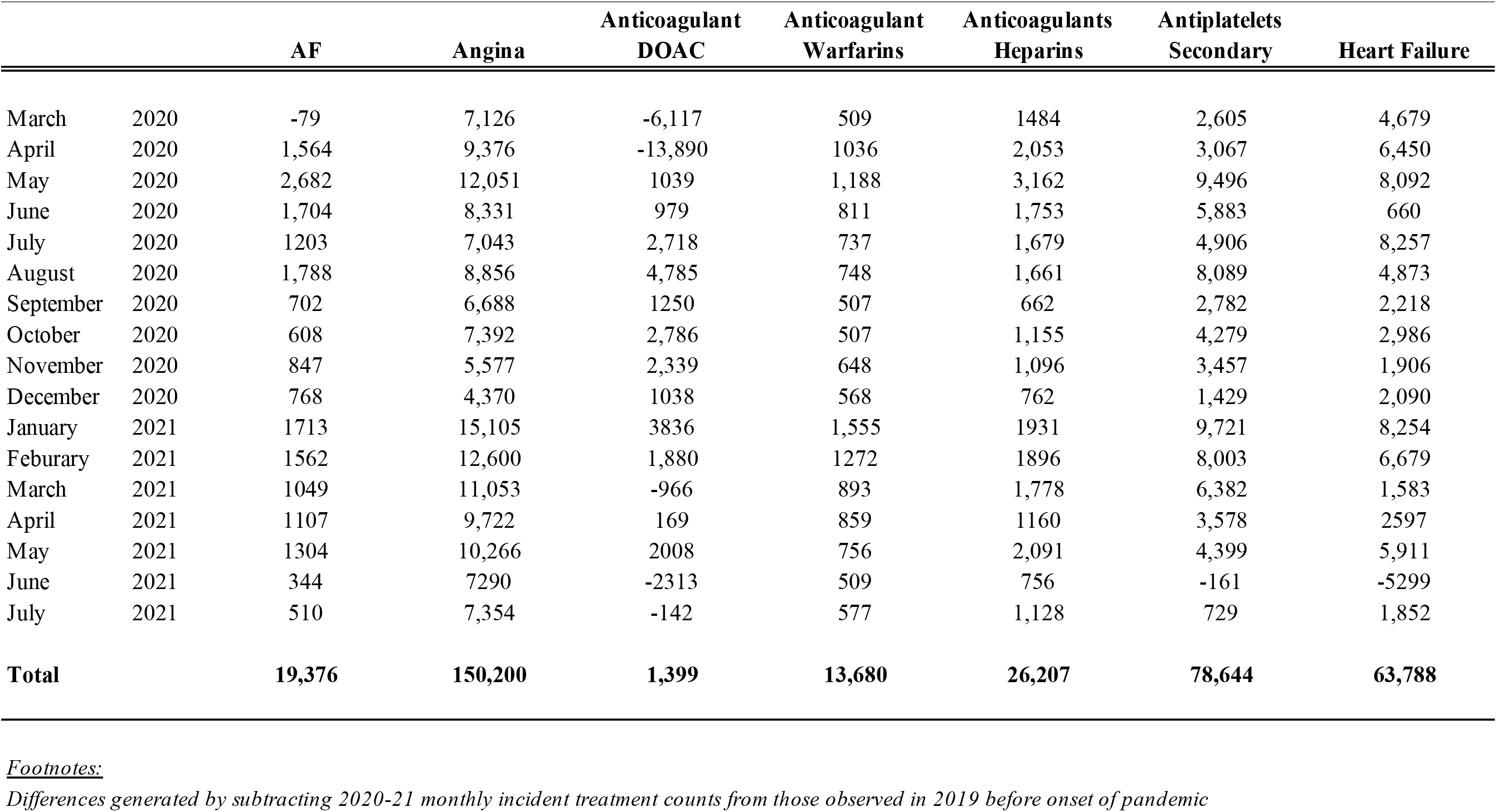
Difference in incident medications dispensed by month by CVD/ CVD risk factor sub-group; 2020-21 versus 2019; England, Scotland and Wales.

**SUPPLEMENTARY TABLE 4:** Number CVD events per 1000 with and without antihypertensive treatment by specified sex and QRISK2%; A) assuming non-treatment ongoing over lifetime, and B) non-treatment duration of five years.

| QRISK2% Treatment effect |  |  | N CVD events per 1000 |  |  |  |  |  |  |
| --- | --- | --- | --- | --- | --- | --- | --- | --- | --- |
|  |  |  | Stable Angina | Unstable Angina | MI | Transient Ischaemic Attack | Stroke | Heart Failure | Total CVD events |
| A) Lifetime |  |  |  |  |  |  |  |  |  |
| male | 11.3 | NT | 100 | 33 | 74 | 29 | 103 | 77 | 416 |
|  |  | Tx | 90 | 30 | 67 | 27 | 98 | 75 | 387 |
|  |  | Additional cases pandemic (NT-Tx) | 10 | 3 | 7 | 2 | 5 | 2 | 29 |
| female | 4.9 | NT | 55 | 14 | 24 | 25 | 94 | 46 | 258 |
|  |  | Tx | 48 | 12 | 21 | 22 | 85 | 43 | 231 |
|  |  | Additional cases pandemic (NT-Tx) | 7 | 2 | 3 | 3 | 9 | 3 | 27 |
| B) 5 years |  |  |  |  |  |  |  |  |  |
| male | 11.3 | NT | 16 | 6 | 16 | 3 | 7 | 4 | 52 |
|  |  | Tx | 14 | 5 | 13 | 3 | 6 | 3 | 44 |
|  |  | Additional cases pandemic (NT-Tx) | 2 | 1 | 3 | 0 | 1 | 1 | 8 |
| female | 4.9 | NT | 7 | 3 | 2 | 4 | 5 | 1 | 22 |
|  |  | Tx | 6 | 2 | 2 | 3 | 4 | 1 | 18 |
|  |  | Additional cases pandemic (NT-Tx) | 1 | 0 | 0 | 1 | 1 | 0 | 4 |

**SUPPLEMENTARY TABLE 5:** Mean quantity of Atorvastatin 40mg and Simvastatin 40mg per dispense by year; England, Scotland and Wales.

| Year | ENGLAND |  |  | SCOTLAND |  |  | WALES |  |  |
| --- | --- | --- | --- | --- | --- | --- | --- | --- | --- |
|  | N dispensed items | N total quantity | Mean annual quantity | N dispensed items | N total quantity | Mean annual quantity | N dispensed items | N total quantity | Mean annual quantity |
| *Atorvastatin 40mg* |  |  |  |  |  |  |  |  |  |
| March 2018-2019 | 10,294,244 | 322,054,501 | 31.3 | 746,156 | 33,989,473 | 45.6 | 677,628 | 19,075,547 | 28.2 |
| March 2019-2020 | 12,376,964 | 387,419,631 | 31.3 | 807,197 | 36,496,001 | 45.2 | 735,329 | 20,688,593 | 28.1 |
| March 2020-2021 | 13,561,221 | 425,599,560 | 31.4 | 861,402 | 39,193,761 | 45.5 | 750,963 | 21,171,855 | 28.2 |
| *Simvastatin 40mg* |  |  |  |  |  |  |  |  |  |
| March 2018-2019 | 10,827,331 | 356,029,047 | 32.9 | 1,385,701 | 63,936,350 | 46.1 | 1,045,131 | 30,456,361 | 29.1 |
| March 2019-2020 | 10,762,415 | 353,679,747 | 32.9 | 1,269,292 | 58,317,033 | 45.9 | 953,478 | 27,777,096 | 29.1 |
| March 2020-2021 | 10,058,333 | 331,826,015 | 33.0 | 1,201,965 | 55,573,536 | 46.2 | 840,153 | 24,555,044 | 29.2 |

*No dispensed data are available in English TRE for March 2018*

## SUPPLEMENTARY FIGURES

**SUPPLEMENTARY FIGURE 1:**
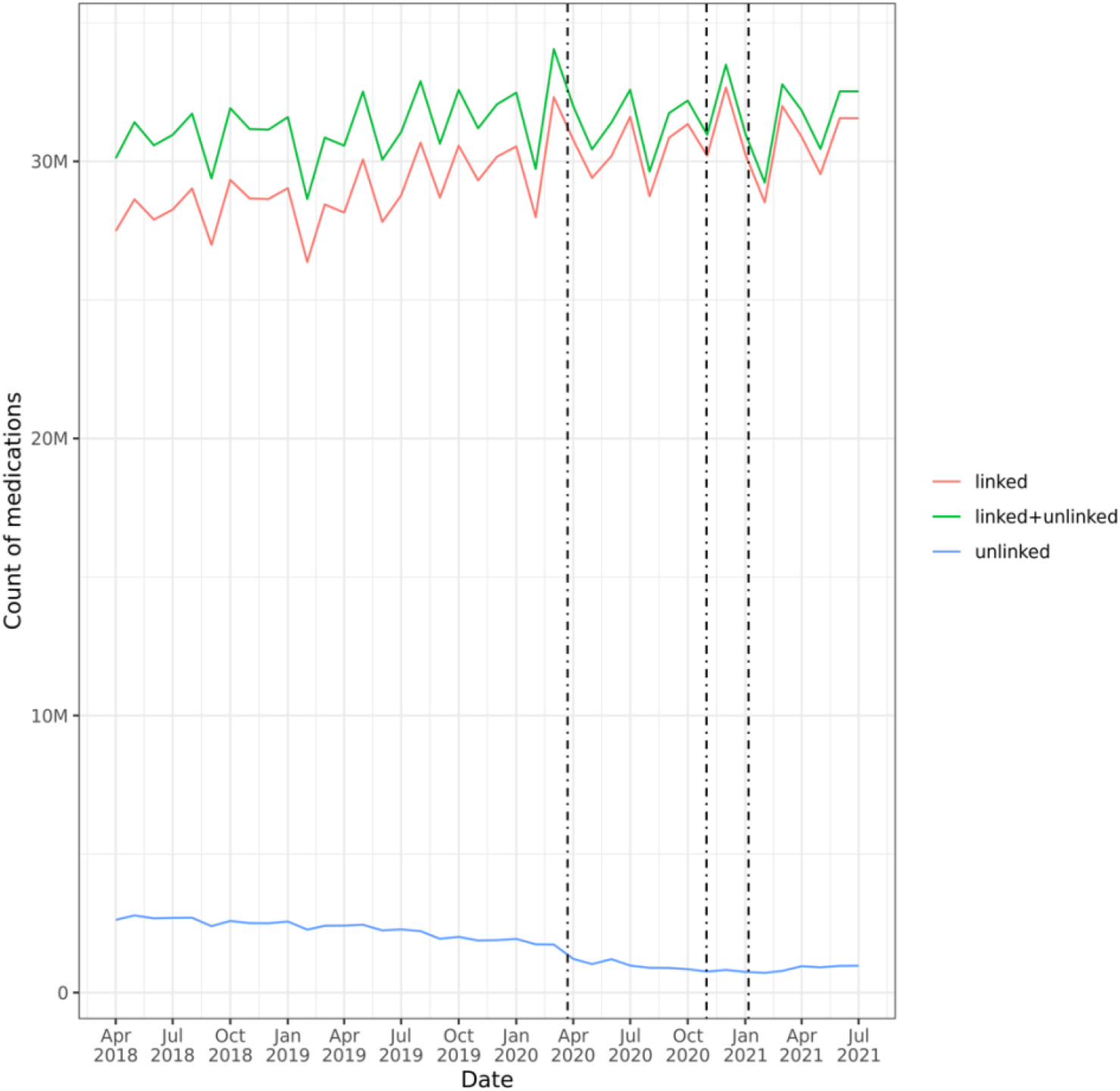
Comparison of trends in counts by month of dispensed CVD medicines, by linkage status. <u>Footnote</u>: “Unlinked” category includes medicines dispensed with invalid (“null”) ID. Null IDs may arise from a variety of reasons, including, but not limited to: problems scanning the NHS number from paper prescriptions, non-availability of date of birth for data linkage, deliberate removal of NHS ID from some sensitive medication. Counts restricted to medicines dispensed to those aged between 18 and 112 years at time of dispense..

**SUPPLEMENTARY FIGURE 2:**
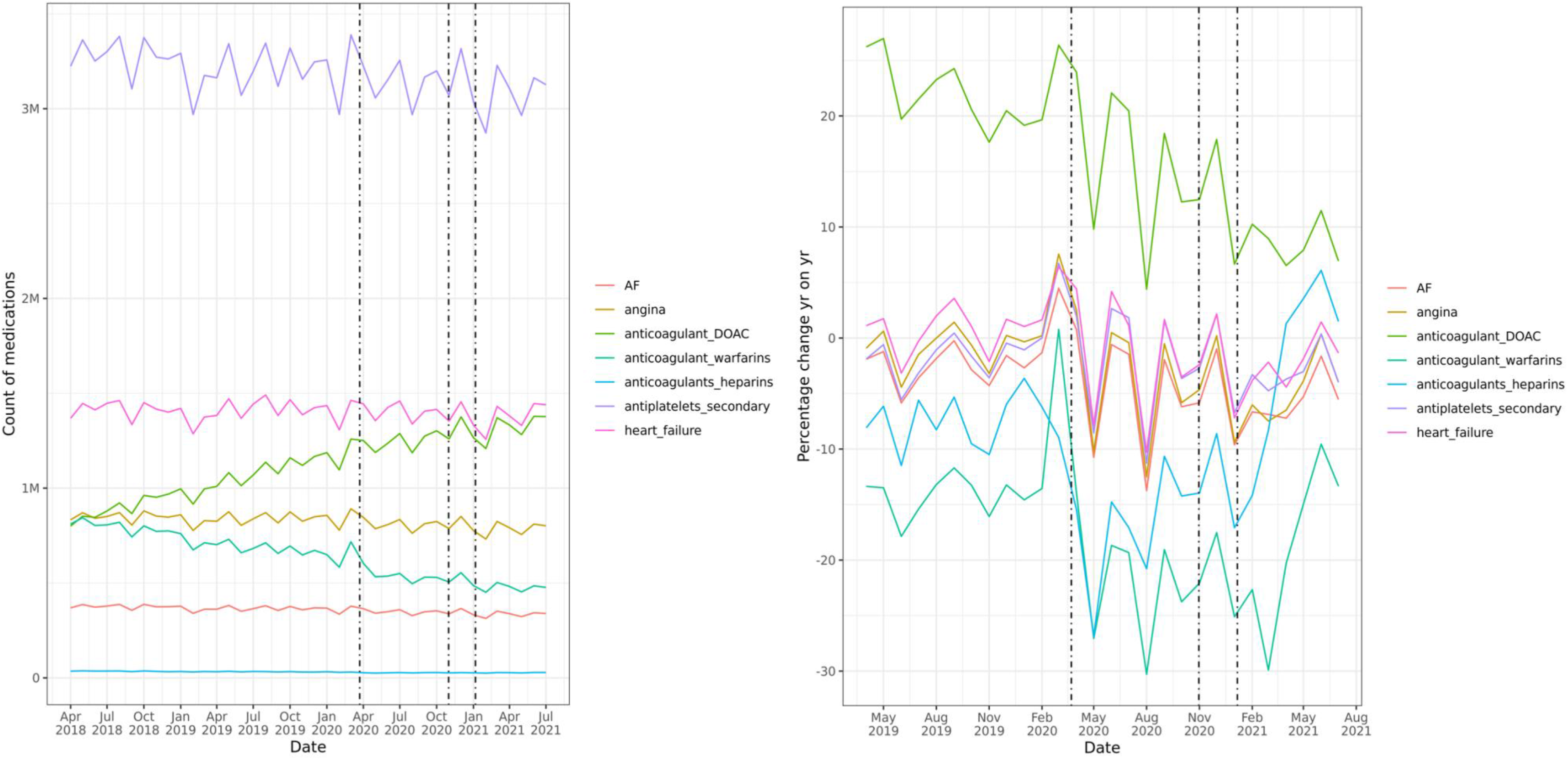
Trends in dispensed CVD medicines over course of pandemic by CVD/ CVD risk factor sub-groups; i) counts by month, and ii) percentage change year-on-year by month.

**SUPPLEMENTARY FIGURE 3a:**
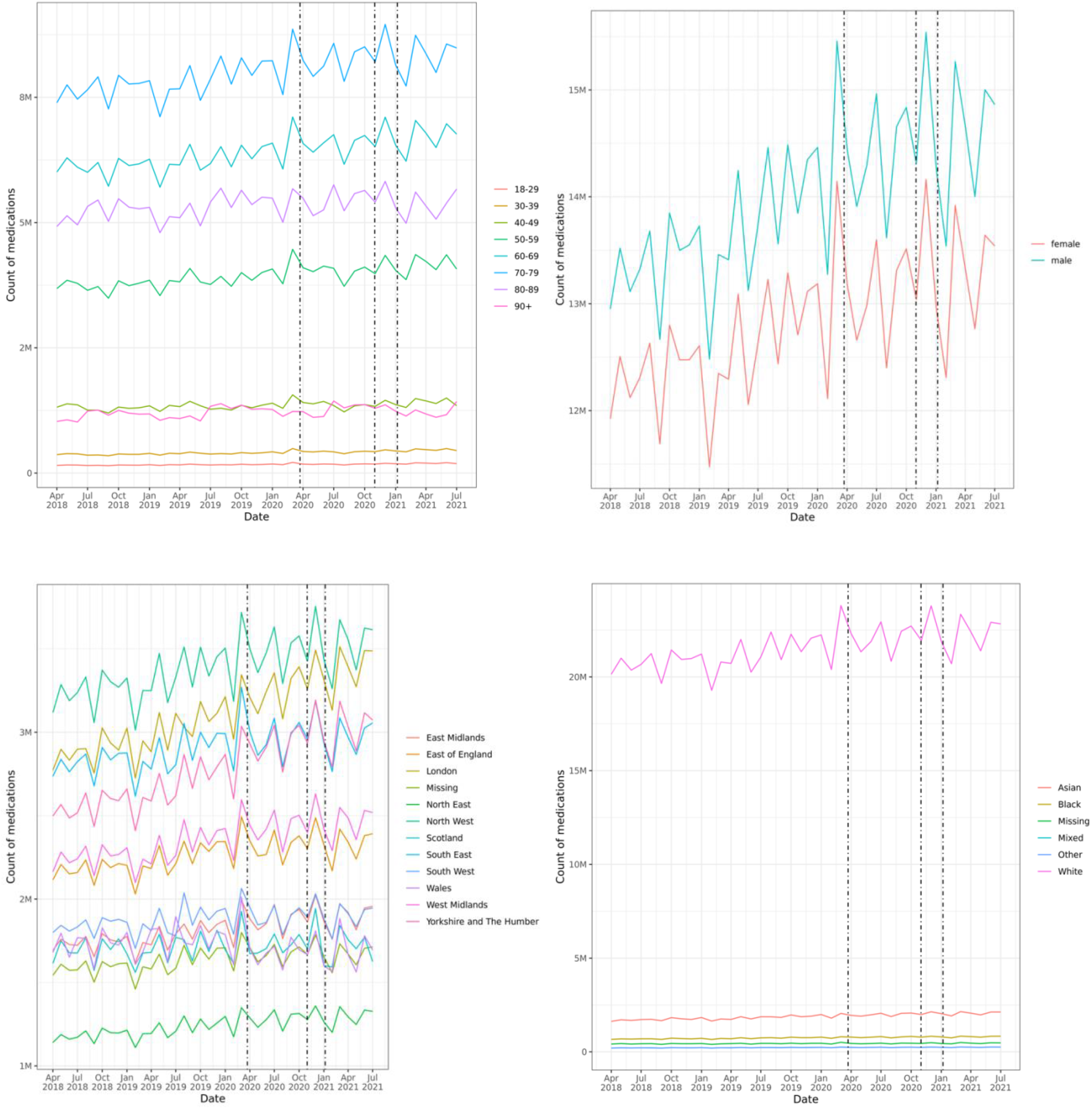
Trends in monthly counts of dispensed CVD medicines (antihypertensives, lipid-lowering medications, T2DM and insulin) stratified by: age, sex, region, ethnicity; England, Scotland and Wales. <u>Footnotes</u>: Dotted lines indicate timing of first, second and third national lockdowns in England. Ethnicity figure includes only English & Welsh data

**SUPPLEMENTARY FIGURE 3b:**
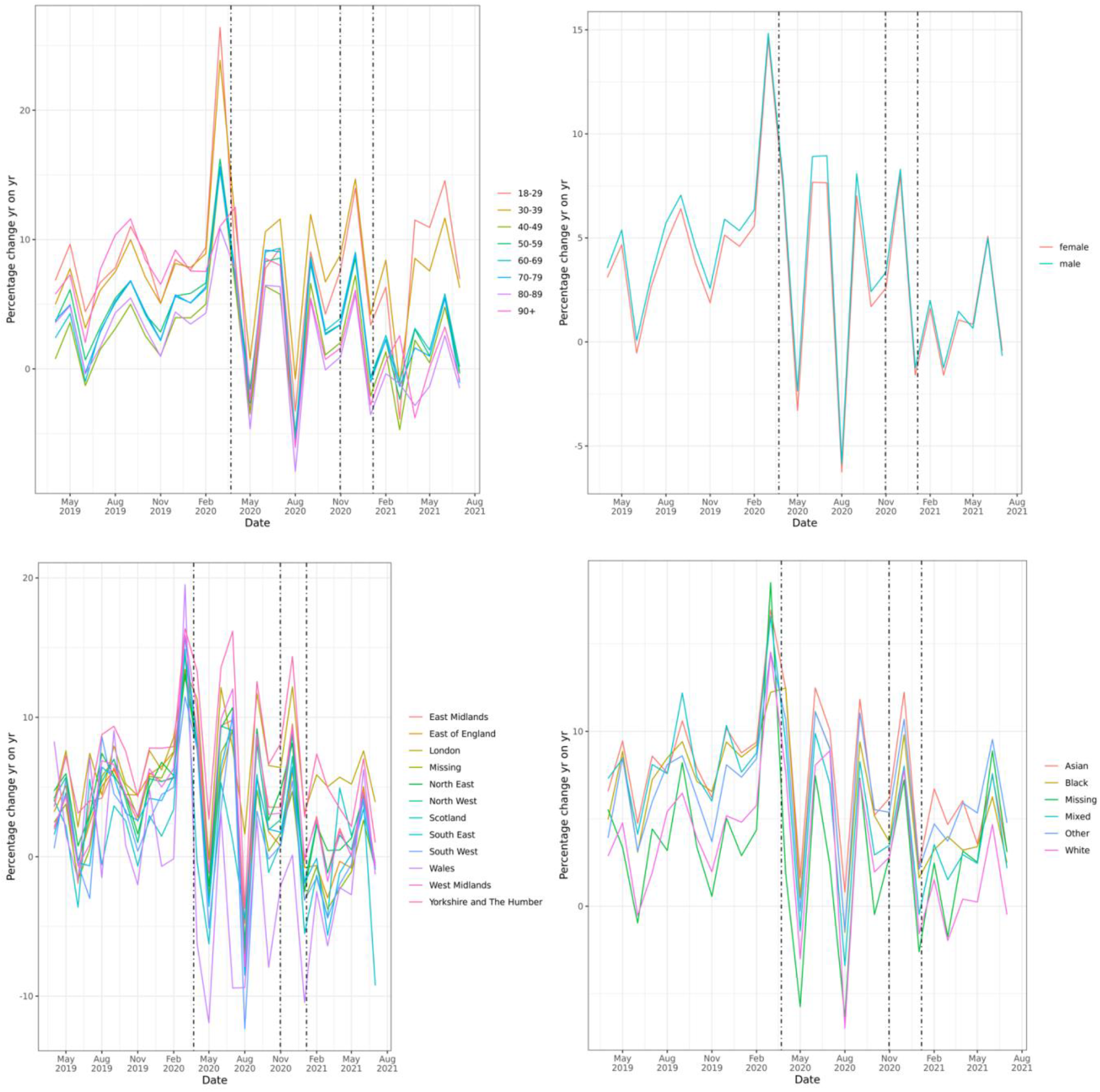
Trends in monthly percentage change year-on-year of dispensed CVD medicines (antihypertensives, lipid-lowering medications, T2DM and insulin) stratified by: age, sex, region, ethnicity. <u>Footnotes</u>: Dotted lines indicate timing of first, second and third national lockdowns in England Ethnicity figure includes only English & Welsh data

**SUPPLEMENTARY FIGURE 3c:**
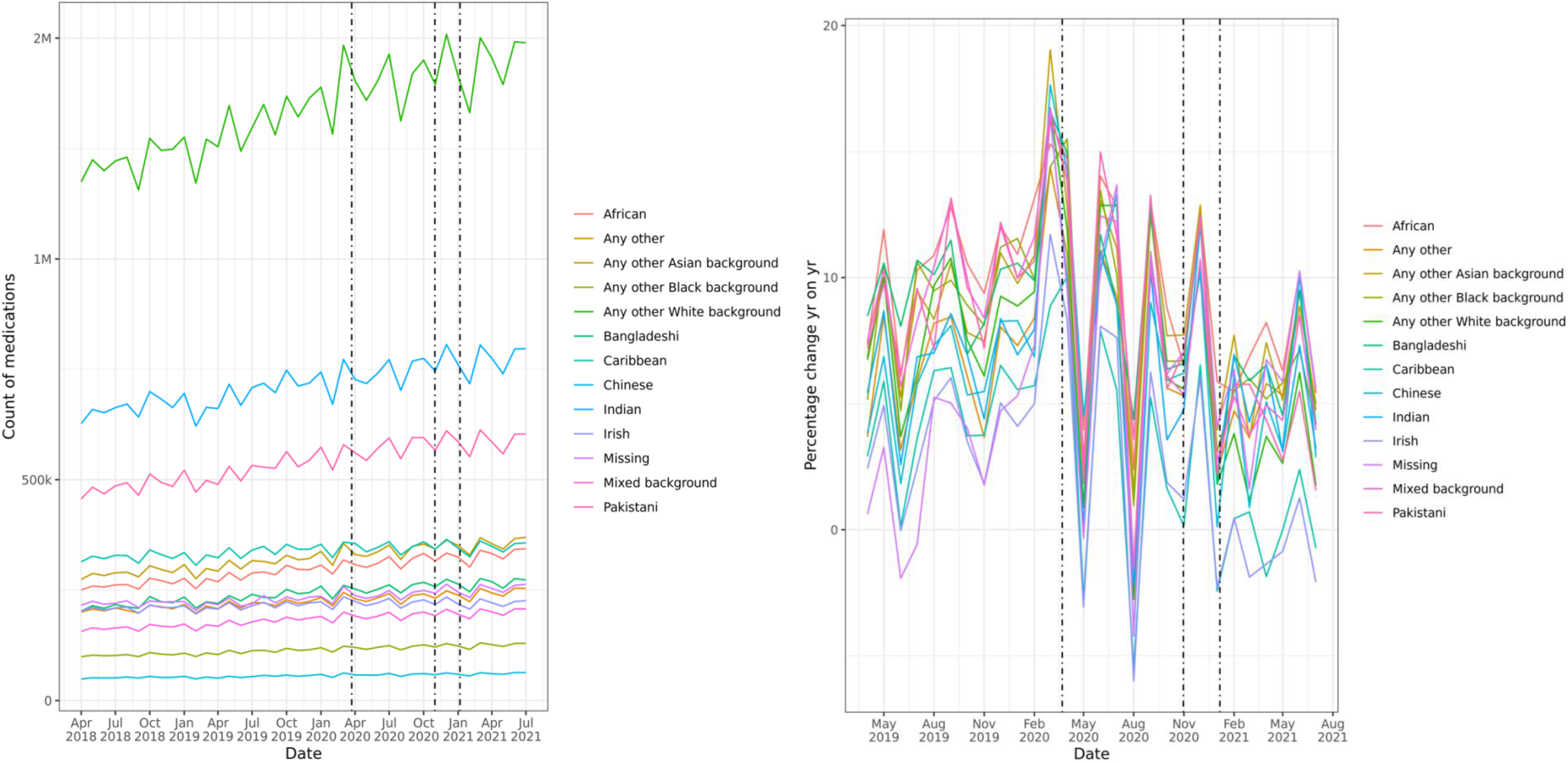
Trends in monthly counts of dispensed CVD medicines (antihypertensives, lipid-lowering medications, T2DM and insulin) detailed ethnicity break-down; England only. <u>Footnote</u>: excludes “British” ethnic group due to scale considerations

**SUPPLEMENTARY FIGURE 4a:**
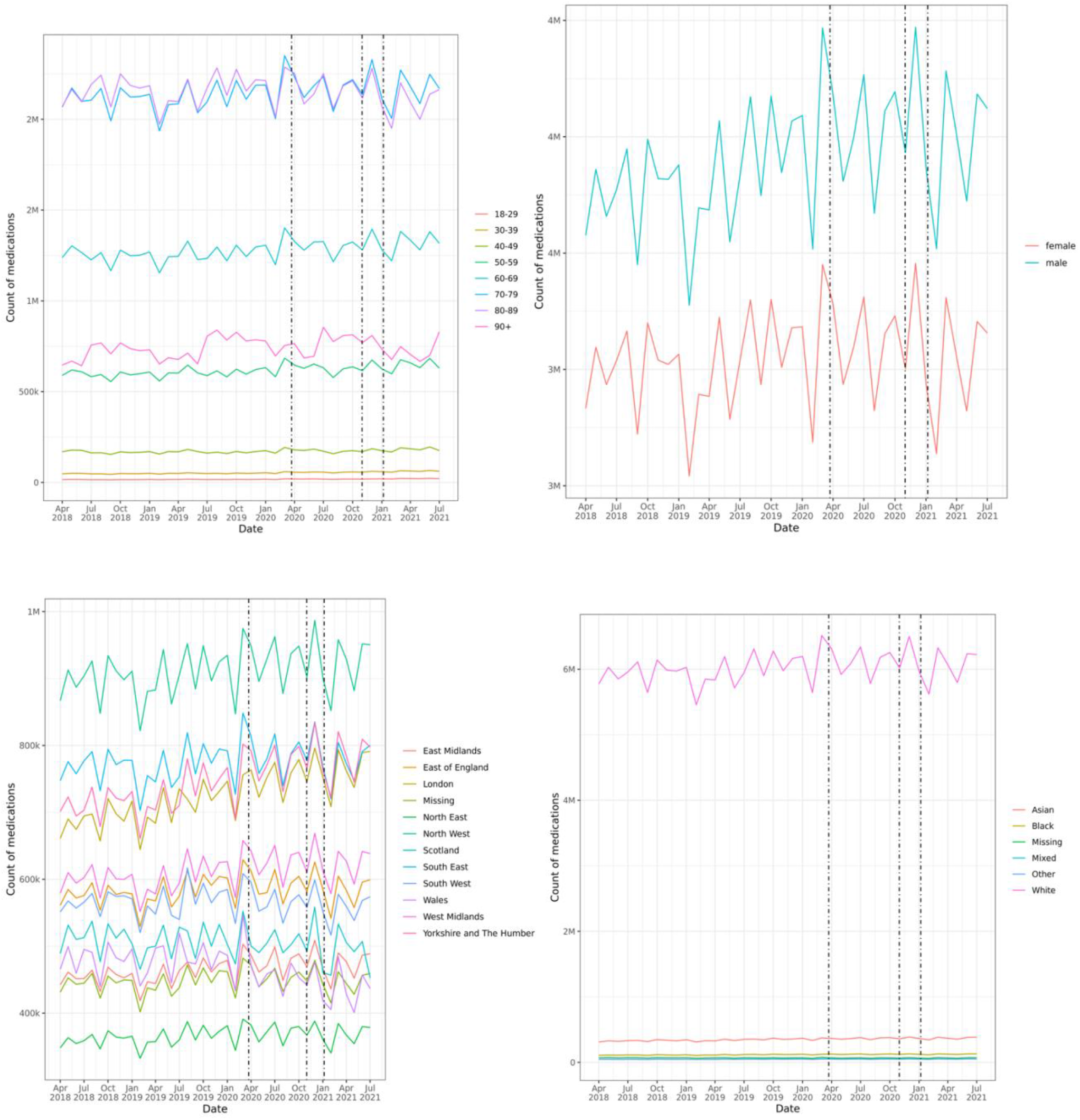
Trends in monthly counts of dispensed CVD medicines (7 sub-groups) stratified by: age, sex, region, ethnicity. <u>Footnotes:</u> Dotted lines indicate timing of first, second and third national lockdowns in England. Ethnicity figure excludes “British” ethnic group due to scale considerations

**SUPPLEMENTARY FIGURE 4b:**
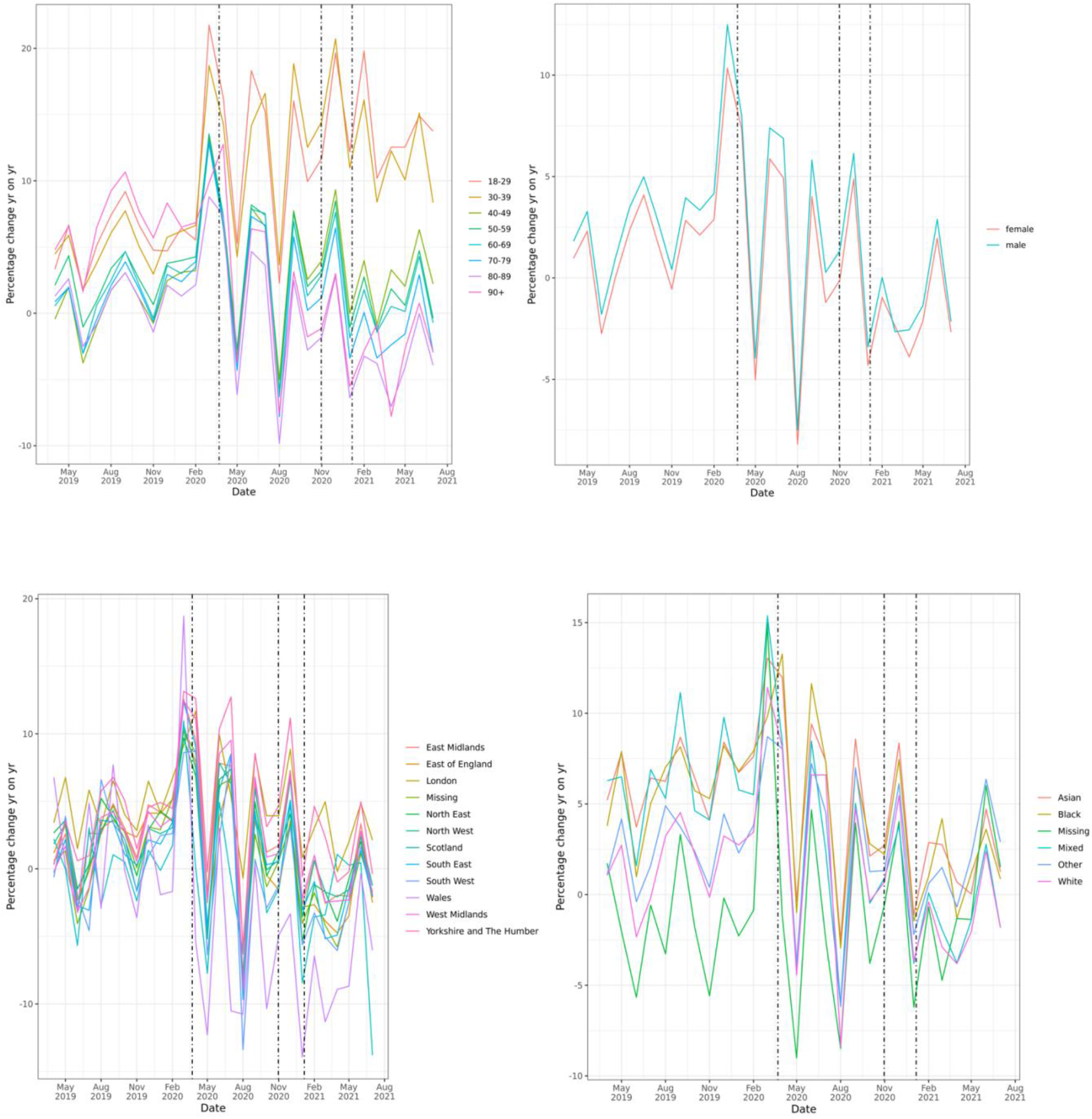
Trends in monthly percentage change year-on-year of dispensed CVD medicines (7 sub-groups) stratified by: age, sex, region, ethnicity. <u>Footnotes:</u> Dotted lines indicate timing of first, second and third national lockdowns in England

**SUPPLEMENTARY FIGURE 5a:**
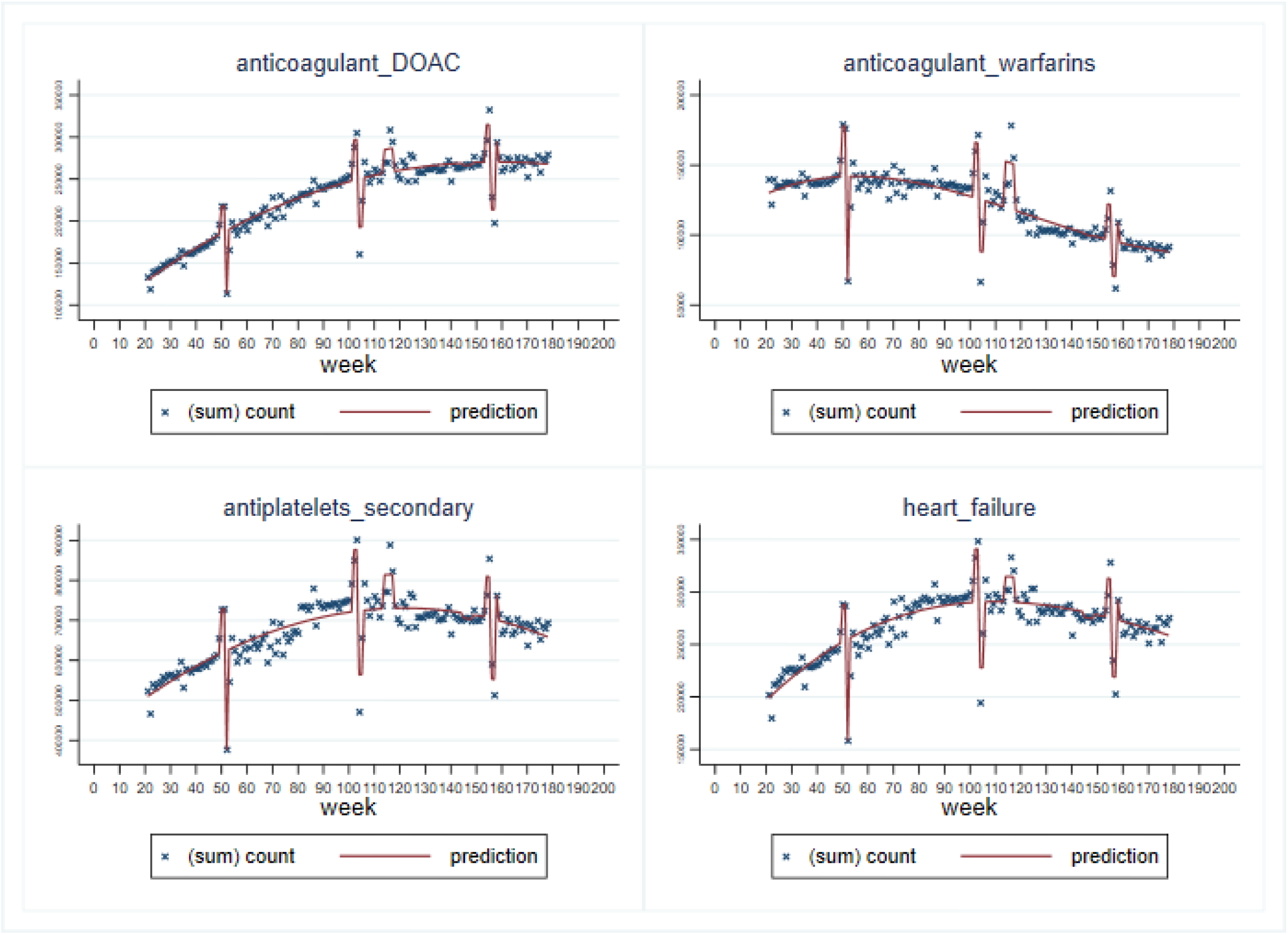
Plot of interrupted time series analysis of prescribed CVD medicines (weekly counts) in England April 2018 to May 2021, by CVD risk factor. <u>Footnotes:</u> Prescribed medicines are analysed for weekly counts; therefore some CVD sub-groups are not available (Angina, Anticoagulants heparins & Atrial Fibrillation). Lower counts at weeks 52, 104-5, 154-5 correspond to Christmas & New Year period 2018, 2019, 2020. Peak at weeks 114-117 corresponds to first national lockdown in England.

**SUPPLEMENTARY FIGURE 5b:**
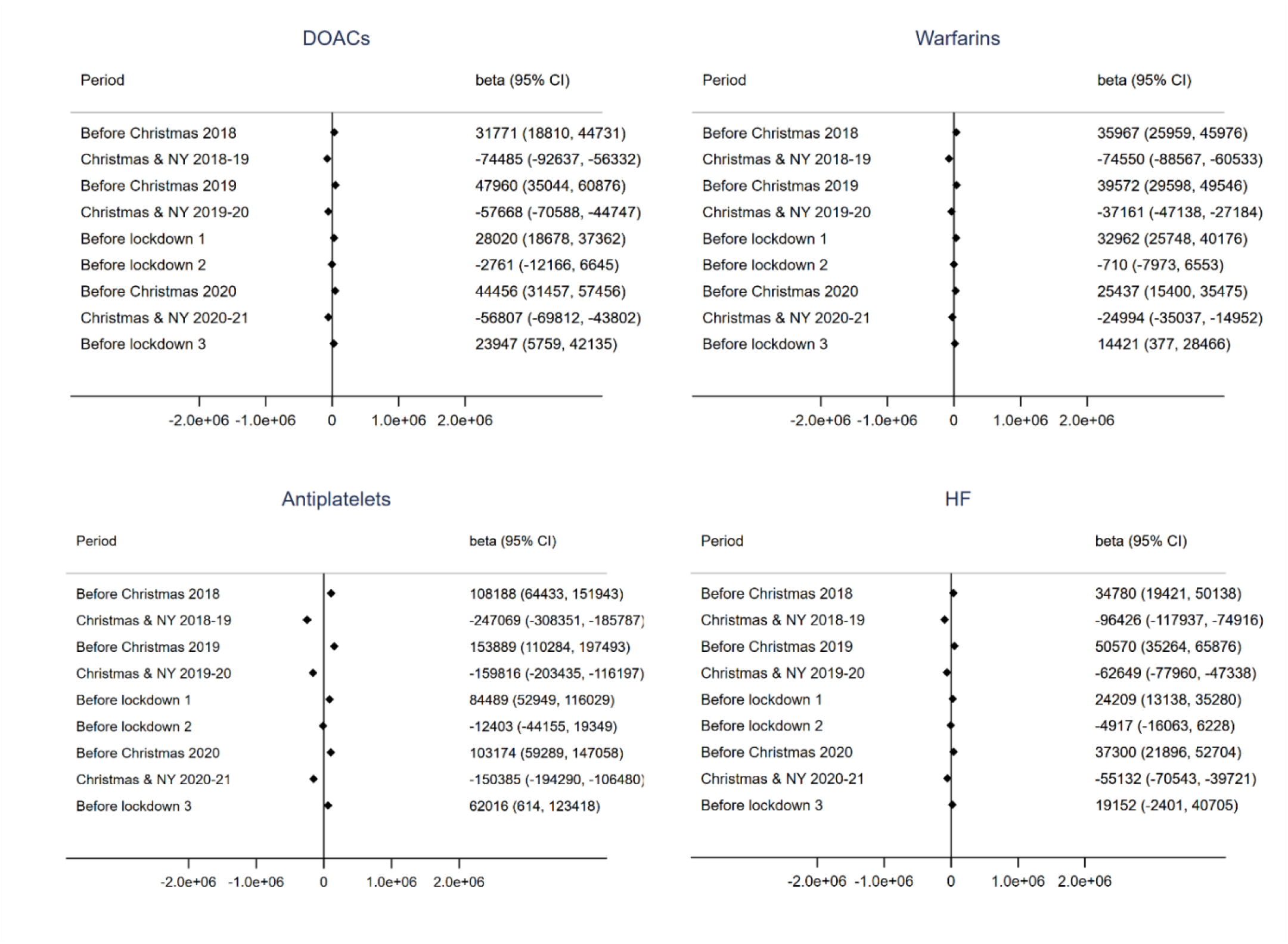
Forest plots of interrupted time series analysis coefficients; prescribed medicines April 2018 to May 2021, by CVD risk factor. <u>Footnote</u>: Beta coefficients (& 95% CIs) of change in count of prescribed medicines associated with specified time period. In the interrupted time series analysis, we additionally adjusted for t, t^*2*^ and t^*3*^

**SUPPLEMENTARY FIGURE 6:**
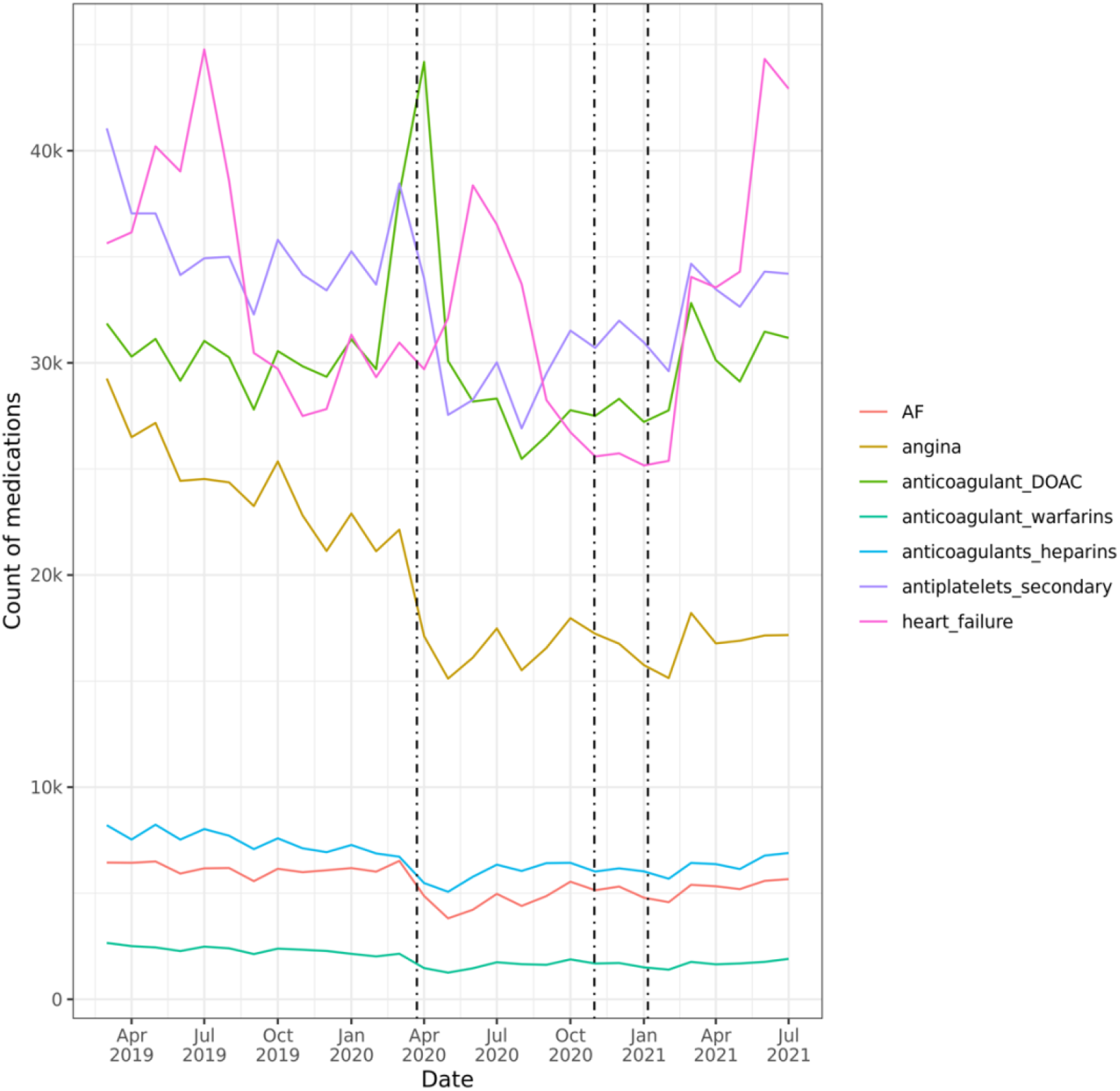
Trends in incident medications dispensed by CVD/ CVD risk factor sub-group; counts by month.

**APPENDIX TABLE 1:**
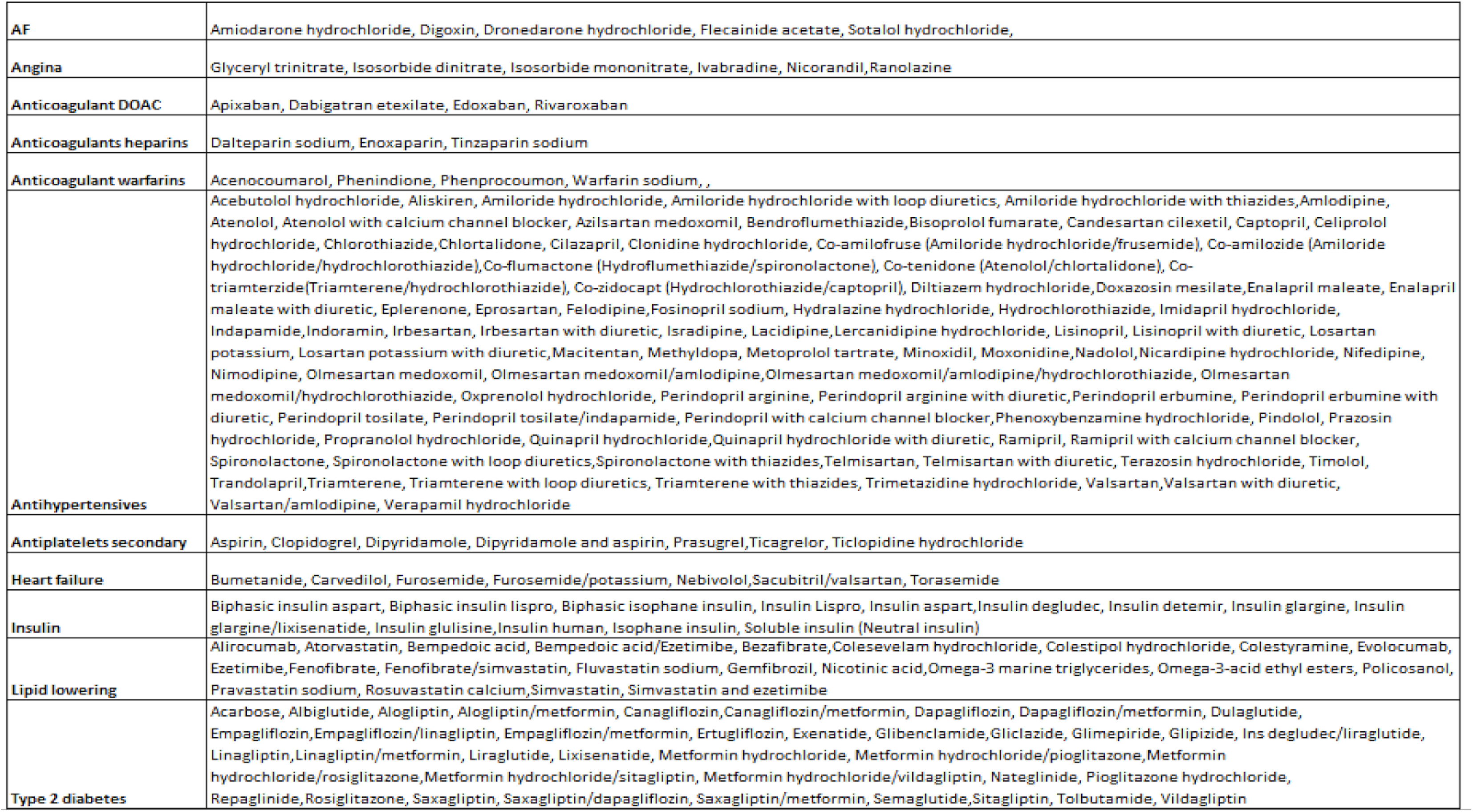
Categorisation of medications (according to drug substance) by CVD/ CVD risk factor category.

